## Supplementary Information for "Using genetic variants to evaluate the causal effect of cholesterol lowering on head and neck cancer risk: a Mendelian randomization study"

**S1 Table. Detailed summary of genetic variants proxying circulating LDL-C, HDL-C, total triglyceride, total cholesterol, apolipoprotein A and B levels.** EA, effect allele or low-density lipoprotein-cholesterol (LDL-C) lowering allele; OA, other or non-effect allele; EAF, effect allele frequency; se, standard error. Beta represents the change in LDL-C levels per copy of the effect allele.

**S2.1 – 2.5 Tables.** **Genetic correlation results for HMGCR, NPC1L1, CETP, PCSK9 and LDLR single nucleotide polymorphisms.** Abbreviations: se, standard error; OR, odds ratio; CIL, lower confidence interval; CIU, upper confidence interval

**S3 Table. Mendelian randomization results of genetically-proxied inhibition of HMGCR, NPC1L1, CETP, PCSK9 and LDLR with risk of combined oral/ oropharyngeal cancer accounting for LD structure in GAME-ON.**

**S4 Table. Assessing weak instrument bias (F-statistic) and proportion of variance in the phenotype (R^2^) explained by the genetic instruments.**

**S5 Table.** **Assessing heterogeneity of single nucleotide polymorphism effect estimates in inverse-variance weighted (IVW) and MR Egger regression for primary analysis.** Abbreviations: Q, Q-statistic; df, degrees of freedom; P, p-value.

**S6 Table. Assessing heterogeneity of single nucleotide polymorphism effect estimates in inverse-variance weighted (IVW) and MR Egger regression for secondary analysis.** Abbreviations: Q, Q-statistic; df, degrees of freedom; P, p-value.

**S7 Table. Assessing directional pleiotropy through MR Egger intercept for primary analysis.** Abbreviations: UVMR, univariable Mendelian randomization; SE, standard error; P, p-value.

**S8 Table. Assessing directional pleiotropy through MR Egger intercept for secondary analysis.** Abbreviations: UVMR, univariable Mendelian randomization; SE, standard error; P, p-value.

**S9 Table. MR-PRESSO results for HMGCR, NPC1L1, CETP, PCSK9, LDLR and other lipid trait SNPs on combined oral/ oropharyngeal cancer.** Abbreviations: RSSobs, residual sum of squares observations.

**S10 Table. Assessing violation of the “NO Measurement Error” (NOME) assumption for instruments used in MR-Egger regression.** Abbreviations: I^2^, I-squared statistic.

**S11 Table. SIMEX correction MR Egger regression results for HMGCR, NPC1L1 and CETP (where I^2^ <0.90).** Abbreviations: OR, odds ratio; CIL, lower confidence interval; CIU, upper confidence interval; P, p-value.

**S12 Table. Mendelian randomization results of genetically-proxied inhibition of HMGCR, NPC1L1, CETP, PCSK9 and LDLR with risk of oral and oropharyngeal cancer including sensitivity analyses in UK Biobank.** Abbreviations: IVW, inverse variance weighted; OR, odds ratio; CI, confidence intervals; P, p-value. OR represents the exponential change in odds of oral/ oropharyngeal squamous cell carcinoma per genetically-proxied inhibition of drug target equivalent to a 1 mmol/L decrease in LDL-C.

**S1 Fig. Forest plots showing Mendelian randomization results for genetically-proxied inhibition of HMGCR, NPC1L1, CETP, PCSK9 and LDLR with risk of combined oral/ oropharyngeal cancer in GAME-ON.**

**S2 Fig. Forest plots showing Mendelian randomization results for LDL-C, HDL-C, total cholesterol, total triglycerides, Apolipoprotein A and Apoprotein B single nucleotide polymorphisms effect on combined oral/oropharyngeal cancer in GAME-ON.**

**S3 Fig. Scatter plots for HMGCR, NPC1L1 and CETP single nucleotide polymorphisms effect on combined oral/oropharyngeal cancer in GAME-ON.**

**S4 Fig. Leave one out analysis for HMGCR, NPC1L1, CETP, PCSK9 and LDLR single nucleotide polymorphisms effect on combined oral/ oropharyngeal cancer in GAME-ON.**

**S5 Fig. Scatter plots for LDL-C, HDL-C, total cholesterol, total triglycerides, apolipoprotein A and apolipoprotein B single nucleotide polymorphisms effect on combined oral/oropharyngeal cancer in GAME-ON.**

**S6 Fig. Leave one out analysis for LDL-C, HDL-C, total cholesterol, total triglycerides, Apolipoprotein A and Apoprotein B single nucleotide polymorphisms effect on combined oral/oropharyngeal cancer in GAME-ON.**

**S7 Fig. Forest plots showing meta-analysed causal effects of cholesterol-lowering HMGCR, NPC1L1, CETP, LDLR and PCSK9 single nucleotide polymorphisms on combined head and neck cancer in UK Biobank and GAME-ON.**

**S1 Table.** Detailed summary of genetic variants proxying circulating LDL-C, HDL-C, total triglyceride, total cholesterol, apolipoprotein A and B levels.

| **Target** | **SNP** | **EA** | **OA** | **Beta** | **se** | **P-value** |
| --- | --- | --- | --- | --- | --- | --- |
| LDL cholesterol | rs7534572* | G | C | 0.0407 | 0.0058 | 1.29E-11 |
| LDL cholesterol | rs2642438 | G | A | 0.0352 | 0.0042 | 7.32E-16 |
| LDL cholesterol | rs2587534 | A | G | 0.0391 | 0.0037 | 8.06E-25 |
| LDL cholesterol | rs17404153 | T | G | -0.0336 | 0.0054 | 1.83E-09 |
| LDL cholesterol | rs10947332 | A | G | 0.0504 | 0.0056 | 6.97E-18 |
| LDL cholesterol | rs6909746 | T | C | -0.0263 | 0.0037 | 7.86E-11 |
| LDL cholesterol | rs2737252 | A | G | -0.0314 | 0.0041 | 7.04E-14 |
| LDL cholesterol | rs174583 | T | C | -0.0522 | 0.0038 | 7.00E-41 |
| LDL cholesterol | rs10893499 | A | G | 0.0521 | 0.0053 | 3.86E-21 |
| LDL cholesterol | rs3184504 | C | T | 0.0268 | 0.0038 | 4.20E-12 |
| LDL cholesterol | rs1169288 | C | A | 0.0375 | 0.004 | 6.45E-21 |
| LDL cholesterol | rs1801689 | C | A | 0.1028 | 0.0139 | 9.81E-12 |
| LDL cholesterol | rs2228603 | T | C | -0.1040 | 0.0072 | 4.43E-44 |
| LDL cholesterol | rs2328223 | C | A | 0.0299 | 0.005 | 5.63E-09 |
| LDL cholesterol | rs6065311 | C | T | 0.0417 | 0.0036 | 1.66E-30 |
| LDL cholesterol | rs1800961 | T | C | -0.0685 | 0.0106 | 6.03E-10 |
| LDL cholesterol | rs12066643 | T | C | -0.0389 | 0.0064 | 1.06E-08 |
| LDL cholesterol | rs4970712 | C | A | 0.0339 | 0.0044 | 2.46E-13 |
| LDL cholesterol | rs72902576 | G | T | -0.0933 | 0.0133 | 9.58E-12 |
| LDL cholesterol | rs12916 | C | T | 0.0733 | 0.0038 | 7.79E-78 |
| LDL cholesterol | rs3757354 | T | C | -0.0382 | 0.0044 | 2.09E-17 |
| LDL cholesterol | rs1408272 | G | T | -0.0520 | 0.0083 | 3.68E-09 |
| LDL cholesterol | rs2390536 | A | G | 0.0223 | 0.0038 | 2.04E-08 |
| LDL cholesterol | rs4722551 | C | T | 0.0391 | 0.0049 | 3.95E-14 |
| LDL cholesterol | rs3780181 | G | A | -0.0445 | 0.0074 | 1.76E-09 |
| LDL cholesterol | rs2419604 | G | A | -0.0302 | 0.004 | 7.49E-14 |
| LDL cholesterol | rs314253 | C | T | -0.0242 | 0.0038 | 3.44E-10 |
| LDL cholesterol | rs6511720 | T | G | -0.2209 | 0.0061 | 1.00E-200 |
| LDL cholesterol | rs2965157 | C | T | -0.1886 | 0.0112 | 7.29E-62 |
| LDL cholesterol | rs7254892 | A | G | -0.4853 | 0.0119 | 1.00E-200 |
| LDL cholesterol | rs676388 | C | T | 0.0265 | 0.0039 | 1.31E-11 |
| LDL cholesterol | rs364585 | G | A | 0.0249 | 0.0038 | 4.28E-10 |
| LDL cholesterol | rs10903129 | G | A | 0.0328 | 0.0037 | 3.03E-17 |
| LDL cholesterol | rs11591147 | T | G | -0.4970 | 0.018 | 8.57E-143 |
| LDL cholesterol | rs6693893 | C | T | -0.0767 | 0.0132 | 2.88E-08 |
| LDL cholesterol | rs2030746 | T | C | 0.0214 | 0.0038 | 8.60E-09 |
| LDL cholesterol | rs16831243 | T | C | 0.0378 | 0.0055 | 9.06E-12 |
| LDL cholesterol | rs1250229 | C | T | 0.0243 | 0.0042 | 3.13E-08 |
| LDL cholesterol | rs7640978 | T | C | -0.0392 | 0.0069 | 9.84E-09 |
| LDL cholesterol | rs4530754 | A | G | 0.0275 | 0.0036 | 3.58E-12 |
| LDL cholesterol | rs112201728 | T | C | 0.0675 | 0.0104 | 8.51E-10 |
| LDL cholesterol | rs1564348 | C | T | 0.0481 | 0.005 | 2.76E-21 |
| LDL cholesterol | rs7832643 | T | G | 0.0339 | 0.0038 | 2.67E-17 |
| LDL cholesterol | rs8017377 | A | G | 0.0303 | 0.0038 | 2.52E-15 |
| LDL cholesterol | rs2738459 | C | A | -0.0532 | 0.0058 | 2.26E-19 |
| LDL cholesterol | rs75687619 | T | G | 0.1735 | 0.0161 | 8.05E-24 |
| LDL cholesterol | rs4253776 | G | A | 0.0311 | 0.0059 | 3.35E-08 |
| LDL cholesterol | rs12748152 | T | C | 0.0499 | 0.0066 | 3.21E-12 |
| LDL cholesterol | rs2495495 | C | T | -0.0342 | 0.0059 | 3.52E-08 |
| LDL cholesterol | rs7551981 | T | G | 0.0472 | 0.0038 | 1.36E-33 |
| LDL cholesterol | rs4970834 | T | C | -0.1503 | 0.0047 | 1.00E-200 |
| LDL cholesterol | rs267733 | G | A | -0.0331 | 0.0053 | 5.29E-09 |
| LDL cholesterol | rs1367117 | A | G | 0.1186 | 0.004 | 9.48E-183 |
| LDL cholesterol | rs6544713 | C | T | -0.0806 | 0.0041 | 4.84E-83 |
| LDL cholesterol | rs13206249 | A | G | -0.0378 | 0.0062 | 4.53E-08 |
| LDL cholesterol | rs16891156 | C | A | 0.0965 | 0.0171 | 8.23E-09 |
| LDL cholesterol | rs2073547 | G | A | 0.0485 | 0.0049 | 1.92E-21 |
| LDL cholesterol | rs13277801 | T | C | -0.0338 | 0.0038 | 3.99E-17 |
| LDL cholesterol | rs1883025 | T | C | -0.0296 | 0.0044 | 6.14E-11 |
| LDL cholesterol | rs579459 | C | T | 0.0665 | 0.0045 | 2.42E-44 |
| LDL cholesterol | rs10832962 | T | C | 0.0320 | 0.004 | 6.62E-14 |
| LDL cholesterol | rs4942486 | C | T | -0.0243 | 0.0037 | 2.26E-11 |
| LDL cholesterol | rs2886232 | C | T | -0.0451 | 0.0064 | 3.88E-11 |
| LDL cholesterol | rs12721109 | A | G | -0.4462 | 0.0183 | 2.99E-122 |
| LDL cholesterol | rs6016373 | G | A | -0.0349 | 0.0037 | 7.95E-19 |
| LDL cholesterol | rs5763662 | T | C | 0.0767 | 0.0121 | 1.19E-08 |
| LDL cholesterol | rs6709904 | G | A | -0.0550 | 0.0085 | 4.58E-10 |
| LDL cholesterol | rs2710642 | A | G | 0.0239 | 0.0038 | 6.09E-09 |
| LDL cholesterol | rs10490626 | A | G | -0.0508 | 0.0069 | 1.70E-12 |
| LDL cholesterol | rs10195252 | C | T | -0.0238 | 0.0039 | 3.81E-08 |
| LDL cholesterol | rs11563251 | T | C | 0.0345 | 0.0062 | 4.50E-08 |
| LDL cholesterol | rs9875338 | A | G | -0.0270 | 0.0037 | 2.21E-11 |
| LDL cholesterol | rs6818397 | G | T | -0.0224 | 0.004 | 1.68E-08 |
| LDL cholesterol | rs6882076 | C | T | 0.0456 | 0.0038 | 3.31E-31 |
| LDL cholesterol | rs2315065 | A | C | 0.1102 | 0.0158 | 5.23E-12 |
| LDL cholesterol | rs9987289 | G | A | 0.0714 | 0.0066 | 8.53E-24 |
| LDL cholesterol | rs2954029* | T | A | -0.0564 | 0.0036 | 2.10E-50 |
| LDL cholesterol | rs964184* | C | G | -0.0855 | 0.0078 | 2.01E-26 |
| LDL cholesterol | rs247616 | T | C | -0.0547 | 0.0041 | 2.57E-37 |
| LDL cholesterol | rs2000999 | A | G | 0.0650 | 0.0046 | 4.22E-41 |
| LDL cholesterol | rs6504872 | T | C | 0.0274 | 0.0037 | 3.48E-13 |
| HDL cholesterol | rs12145743 | G | T | 0.0203 | 0.0036 | 1.80E-08 |
| HDL cholesterol | rs4650994 | A | G | -0.0210 | 0.0034 | 6.70E-09 |
| HDL cholesterol | rs17145738 | T | C | 0.0408 | 0.0053 | 4.95E-13 |
| HDL cholesterol | rs1866956 | T | C | 0.0217 | 0.0037 | 7.96E-10 |
| HDL cholesterol | rs11789603 | T | C | 0.0600 | 0.006 | 3.69E-21 |
| HDL cholesterol | rs964184* | C | G | 0.1065 | 0.0071 | 6.09E-48 |
| HDL cholesterol | rs7112577* | G | C | 0.0826 | 0.0129 | 2.34E-10 |
| HDL cholesterol | rs2454722 | G | A | 0.0351 | 0.0044 | 3.31E-14 |
| HDL cholesterol | rs4379922 | C | T | 0.0247 | 0.0036 | 9.56E-12 |
| HDL cholesterol | rs10468017 | T | C | 0.1179 | 0.0038 | 1.21E-188 |
| HDL cholesterol | rs2278236 | A | G | 0.0331 | 0.0035 | 3.18E-18 |
| HDL cholesterol | rs737337 | C | T | -0.0565 | 0.0061 | 4.56E-17 |
| HDL cholesterol | rs12133576 | G | A | -0.0243 | 0.0035 | 6.15E-11 |
| HDL cholesterol | rs1689797 | A | C | -0.0358 | 0.0036 | 2.85E-21 |
| HDL cholesterol | rs4846914 | A | G | 0.0479 | 0.0034 | 3.51E-41 |
| HDL cholesterol | rs1047891 | A | C | -0.0269 | 0.0039 | 8.73E-10 |
| HDL cholesterol | rs13099479 | A | G | 0.0360 | 0.0062 | 1.82E-08 |
| HDL cholesterol | rs10019888 | G | A | -0.0270 | 0.0046 | 4.90E-08 |
| HDL cholesterol | rs6450176 | A | G | -0.0254 | 0.0039 | 6.87E-10 |
| HDL cholesterol | rs998584 | A | C | -0.0260 | 0.0038 | 2.27E-11 |
| HDL cholesterol | rs1936800* | T | C | -0.0200 | 0.0034 | 3.05E-10 |
| HDL cholesterol | rs4917014 | G | T | 0.0222 | 0.0036 | 1.03E-08 |
| HDL cholesterol | rs12412743 | T | C | -0.0291 | 0.0045 | 1.31E-09 |
| HDL cholesterol | rs3847502 | A | C | 0.0480 | 0.0036 | 3.31E-38 |
| HDL cholesterol | rs102275 | C | T | -0.0391 | 0.0035 | 6.40E-28 |
| HDL cholesterol | rs3741414 | T | C | 0.0296 | 0.004 | 6.10E-14 |
| HDL cholesterol | rs2241770 | C | T | -0.0989 | 0.0057 | 6.78E-60 |
| HDL cholesterol | rs4148005 | G | T | -0.0283 | 0.0036 | 5.74E-14 |
| HDL cholesterol | rs4465830 | G | A | -0.0597 | 0.0044 | 5.17E-40 |
| HDL cholesterol | rs4660293 | G | A | -0.0353 | 0.004 | 2.86E-18 |
| HDL cholesterol | rs12740374 | T | G | 0.0343 | 0.0041 | 1.69E-15 |
| HDL cholesterol | rs2642438 | G | A | 0.0303 | 0.0039 | 7.78E-14 |
| HDL cholesterol | rs1515110 | T | G | -0.0323 | 0.0035 | 8.04E-18 |
| HDL cholesterol | rs2290547 | A | G | -0.0297 | 0.0046 | 3.69E-09 |
| HDL cholesterol | rs2013208 | T | C | 0.0254 | 0.0036 | 8.92E-12 |
| HDL cholesterol | rs6805251 | C | T | -0.0200 | 0.0035 | 1.33E-08 |
| HDL cholesterol | rs13076253 | C | A | -0.0283 | 0.0048 | 4.96E-09 |
| HDL cholesterol | rs3822072 | A | G | -0.0251 | 0.0034 | 4.06E-12 |
| HDL cholesterol | rs2602836 | G | A | -0.0192 | 0.0034 | 4.96E-08 |
| HDL cholesterol | rs9457931 | G | A | -0.0552 | 0.0073 | 7.30E-13 |
| HDL cholesterol | rs4240624 | A | G | 0.0818 | 0.0058 | 1.32E-45 |
| HDL cholesterol | rs2293889 | G | T | 0.0312 | 0.0035 | 4.27E-17 |
| HDL cholesterol | rs10808546 | T | C | 0.0409 | 0.0034 | 4.11E-30 |
| HDL cholesterol | rs1883025 | T | C | -0.0698 | 0.0041 | 1.50E-65 |
| HDL cholesterol | rs10761771 | C | T | 0.0198 | 0.0034 | 4.12E-09 |
| HDL cholesterol | rs499974 | A | C | -0.0263 | 0.0044 | 1.12E-08 |
| HDL cholesterol | rs11045163 | G | A | 0.0217 | 0.0035 | 3.20E-09 |
| HDL cholesterol | rs11065987 | G | A | -0.0222 | 0.0035 | 1.23E-09 |
| HDL cholesterol | rs4983559 | A | G | -0.0197 | 0.0036 | 9.57E-09 |
| HDL cholesterol | rs424346 | T | C | 0.0679 | 0.0113 | 4.84E-08 |
| HDL cholesterol | rs9989419 | G | A | 0.1473 | 0.0036 | 1.00E-200 |
| HDL cholesterol | rs16965220 | A | C | 0.0219 | 0.0037 | 7.91E-09 |
| HDL cholesterol | rs1877031 | A | G | 0.0336 | 0.0036 | 1.20E-19 |
| HDL cholesterol | rs4969178 | G | A | 0.0263 | 0.0035 | 1.53E-12 |
| HDL cholesterol | rs2075650 | G | A | -0.0554 | 0.0051 | 9.72E-26 |
| HDL cholesterol | rs2288912* | G | C | 0.0297 | 0.0036 | 7.15E-15 |
| HDL cholesterol | rs103294 | T | C | 0.0523 | 0.0044 | 3.99E-30 |
| HDL cholesterol | rs181360 | G | T | -0.0376 | 0.0042 | 9.24E-18 |
| HDL cholesterol | rs12748152 | T | C | -0.0506 | 0.0062 | 9.74E-16 |
| HDL cholesterol | rs676210 | A | G | 0.0660 | 0.004 | 2.34E-54 |
| HDL cholesterol | rs7607980 | C | T | 0.0447 | 0.0052 | 1.81E-15 |
| HDL cholesterol | rs687339 | T | C | -0.0316 | 0.0042 | 7.11E-13 |
| HDL cholesterol | rs3861397 | G | A | -0.0240 | 0.0036 | 8.40E-11 |
| HDL cholesterol | rs702485 | G | A | 0.0243 | 0.0034 | 6.45E-12 |
| HDL cholesterol | rs11765979 | C | A | 0.0412 | 0.0048 | 3.11E-17 |
| HDL cholesterol | rs13702 | C | T | 0.1058 | 0.0038 | 1.28E-160 |
| HDL cholesterol | rs10087900 | A | G | -0.0231 | 0.0036 | 2.17E-09 |
| HDL cholesterol | rs2066714 | C | T | 0.0453 | 0.0071 | 7.26E-10 |
| HDL cholesterol | rs970548 | C | A | 0.0258 | 0.0039 | 1.71E-10 |
| HDL cholesterol | rs2241210 | G | A | 0.0332 | 0.0035 | 2.49E-20 |
| HDL cholesterol | rs838876 | G | A | -0.0493 | 0.0039 | 7.32E-33 |
| HDL cholesterol | rs4939883 | C | T | 0.0799 | 0.0045 | 1.80E-66 |
| HDL cholesterol | rs6567160 | C | T | -0.0257 | 0.0041 | 2.92E-09 |
| HDL cholesterol | rs731839 | A | G | 0.0220 | 0.0037 | 3.44E-09 |
| HDL cholesterol | rs6031587 | T | C | -0.0488 | 0.0074 | 1.92E-09 |
| HDL cholesterol | rs2606736 | T | C | -0.0246 | 0.0043 | 4.80E-08 |
| HDL cholesterol | rs13107325 | T | C | -0.0708 | 0.0078 | 1.06E-15 |
| HDL cholesterol | rs1980493 | C | T | -0.0318 | 0.0048 | 3.76E-10 |
| HDL cholesterol | rs205262 | G | A | -0.0283 | 0.0039 | 3.88E-13 |
| HDL cholesterol | rs4142995 | T | G | -0.0263 | 0.0037 | 9.36E-12 |
| HDL cholesterol | rs17173637 | C | T | -0.0363 | 0.0057 | 1.90E-08 |
| HDL cholesterol | rs686030 | A | C | 0.0550 | 0.0049 | 4.29E-27 |
| HDL cholesterol | rs2250802 | A | G | -0.0340 | 0.0038 | 2.02E-17 |
| HDL cholesterol | rs12801636 | A | G | 0.0235 | 0.0042 | 3.15E-08 |
| HDL cholesterol | rs7306660 | A | G | -0.0345 | 0.0036 | 3.34E-19 |
| HDL cholesterol | rs492571 | C | T | -0.0663 | 0.009 | 1.27E-12 |
| HDL cholesterol | rs633695 | G | A | 0.0885 | 0.0054 | 7.82E-58 |
| HDL cholesterol | rs16942887 | A | G | 0.0831 | 0.0051 | 8.28E-54 |
| HDL cholesterol | rs2925979 | C | T | 0.0351 | 0.0037 | 1.32E-19 |
| Total cholesterol | rs11802413 | T | C | 0.0287 | 0.0035 | 1.58E-14 |
| Total cholesterol | rs4988235 | A | G | -0.0308 | 0.004 | 3.97E-14 |
| Total cholesterol | rs11694172 | G | A | 0.0277 | 0.0041 | 1.95E-09 |
| Total cholesterol | rs6818397 | G | T | -0.0254 | 0.0039 | 9.51E-11 |
| Total cholesterol | rs4530754 | A | G | 0.0228 | 0.0035 | 1.68E-09 |
| Total cholesterol | rs2814982 | T | C | -0.0441 | 0.0057 | 3.68E-15 |
| Total cholesterol | rs2737252 | A | G | -0.0331 | 0.0039 | 1.63E-16 |
| Total cholesterol | rs1883025 | T | C | -0.0671 | 0.0042 | 5.75E-53 |
| Total cholesterol | rs1535 | G | A | -0.0497 | 0.0037 | 8.62E-39 |
| Total cholesterol | rs2000999 | A | G | 0.0617 | 0.0044 | 6.80E-41 |
| Total cholesterol | rs6504872 | T | C | 0.0250 | 0.0035 | 6.99E-12 |
| Total cholesterol | rs2738459 | C | A | -0.0387 | 0.0057 | 2.11E-11 |
| Total cholesterol | rs2228603 | T | C | -0.1217 | 0.0069 | 1.05E-62 |
| Total cholesterol | rs281393 | T | C | -0.0322 | 0.0055 | 4.26E-08 |
| Total cholesterol | rs11591147 | T | G | -0.3341 | 0.0173 | 8.83E-86 |
| Total cholesterol | rs646776 | T | C | 0.1272 | 0.0042 | 4.78E-187 |
| Total cholesterol | rs558971 | G | A | 0.0398 | 0.0036 | 7.02E-28 |
| Total cholesterol | rs2030746 | T | C | 0.0199 | 0.0037 | 3.60E-08 |
| Total cholesterol | rs13315871 | A | G | -0.0355 | 0.0061 | 3.48E-08 |
| Total cholesterol | rs3757354 | T | C | -0.0348 | 0.0042 | 2.22E-15 |
| Total cholesterol | rs1997243 | G | A | 0.0332 | 0.005 | 2.72E-10 |
| Total cholesterol | rs581080* | C | G | 0.0377 | 0.0047 | 1.02E-13 |
| Total cholesterol | rs12412743 | T | C | -0.0298 | 0.0047 | 6.98E-10 |
| Total cholesterol | rs4752805 | G | A | 0.0251 | 0.0041 | 1.62E-09 |
| Total cholesterol | rs247616 | T | C | 0.0499 | 0.004 | 4.47E-32 |
| Total cholesterol | rs2886232 | C | T | -0.0358 | 0.0062 | 3.87E-08 |
| Total cholesterol | rs8103315 | A | C | 0.0422 | 0.0055 | 5.94E-15 |
| Total cholesterol | rs75687619 | T | G | 0.1592 | 0.0153 | 3.61E-22 |
| Total cholesterol | rs6016373 | G | A | -0.0319 | 0.0036 | 1.00E-17 |
| Total cholesterol | rs1800961 | T | C | -0.1062 | 0.0101 | 1.34E-24 |
| Total cholesterol | rs181360 | G | T | -0.0278 | 0.0043 | 7.32E-10 |
| Total cholesterol | rs2642438 | G | A | 0.0370 | 0.004 | 1.28E-18 |
| Total cholesterol | rs17526895 | G | A | -0.0420 | 0.0067 | 5.78E-09 |
| Total cholesterol | rs2287623 | A | G | -0.0273 | 0.0036 | 4.09E-12 |
| Total cholesterol | rs12916 | C | T | 0.0684 | 0.0036 | 4.55E-74 |
| Total cholesterol | rs9272775 | C | T | 0.0317 | 0.0055 | 2.13E-08 |
| Total cholesterol | rs12670798 | C | T | 0.0364 | 0.0041 | 9.48E-17 |
| Total cholesterol | rs9987289 | G | A | 0.0842 | 0.0063 | 1.84E-36 |
| Total cholesterol | rs2954029* | T | A | -0.0622 | 0.0035 | 2.42E-65 |
| Total cholesterol | rs7832643 | T | G | 0.0289 | 0.0037 | 3.12E-13 |
| Total cholesterol | rs11789603 | T | C | 0.0427 | 0.0062 | 1.44E-11 |
| Total cholesterol | rs10832962 | T | C | 0.0315 | 0.0039 | 1.54E-14 |
| Total cholesterol | rs964184* | C | G | -0.1214 | 0.0076 | 2.84E-55 |
| Total cholesterol | rs3184504 | C | T | 0.0318 | 0.0037 | 1.62E-17 |
| Total cholesterol | rs10773003 | A | G | 0.0369 | 0.0058 | 4.08E-09 |
| Total cholesterol | rs6511720 | T | G | -0.1851 | 0.0059 | 1.00E-200 |
| Total cholesterol | rs7534572* | G | C | 0.0629 | 0.0055 | 3.60E-28 |
| Total cholesterol | rs6603981 | T | C | 0.0351 | 0.0043 | 7.85E-15 |
| Total cholesterol | rs9306897 | C | T | -0.0488 | 0.0037 | 7.51E-37 |
| Total cholesterol | rs780093 | C | T | -0.0515 | 0.0036 | 2.59E-42 |
| Total cholesterol | rs6544713 | C | T | -0.0773 | 0.004 | 1.69E-81 |
| Total cholesterol | rs7616006 | G | A | -0.0315 | 0.0036 | 8.41E-17 |
| Total cholesterol | rs6882076 | C | T | 0.0508 | 0.0037 | 5.35E-41 |
| Total cholesterol | rs11153594 | T | C | -0.0290 | 0.0036 | 1.27E-14 |
| Total cholesterol | rs2315065 | A | C | 0.1102 | 0.0158 | 1.10E-11 |
| Total cholesterol | rs2073547 | G | A | 0.0456 | 0.0047 | 3.83E-21 |
| Total cholesterol | rs10088180 | G | A | -0.0228 | 0.004 | 6.02E-10 |
| Total cholesterol | rs4738684 | G | A | -0.0392 | 0.0037 | 1.12E-23 |
| Total cholesterol | rs2066714 | C | T | 0.0442 | 0.0076 | 1.14E-08 |
| Total cholesterol | rs579459 | C | T | 0.0620 | 0.0044 | 8.83E-42 |
| Total cholesterol | rs10904908 | G | A | 0.0250 | 0.0036 | 2.60E-11 |
| Total cholesterol | rs2255141 | G | A | -0.0314 | 0.0039 | 6.51E-16 |
| Total cholesterol | rs11220462 | A | G | 0.0474 | 0.0058 | 5.49E-15 |
| Total cholesterol | rs4883201 | G | A | -0.0350 | 0.0056 | 1.74E-09 |
| Total cholesterol | rs2244608 | G | A | 0.0313 | 0.0037 | 9.62E-18 |
| Total cholesterol | rs2156552* | T | A | 0.0570 | 0.0047 | 1.25E-31 |
| Total cholesterol | rs386003 | T | G | 0.0344 | 0.0058 | 4.52E-08 |
| Total cholesterol | rs2277862 | T | C | -0.0349 | 0.0052 | 5.26E-11 |
| Total cholesterol | rs2235367 | G | A | 0.0357 | 0.0035 | 7.22E-25 |
| Total cholesterol | rs138777 | G | A | -0.0214 | 0.0037 | 4.74E-08 |
| Total cholesterol | rs7551981 | T | G | 0.0358 | 0.0037 | 7.50E-22 |
| Total cholesterol | rs6709904 | G | A | -0.0545 | 0.0083 | 8.39E-10 |
| Total cholesterol | rs11563251 | T | C | 0.0368 | 0.0059 | 1.27E-09 |
| Total cholesterol | rs7640978 | T | C | -0.0376 | 0.0066 | 1.66E-08 |
| Total cholesterol | rs1800562 | A | G | -0.0565 | 0.0077 | 1.91E-12 |
| Total cholesterol | rs9391858 | G | A | 0.0495 | 0.005 | 7.20E-22 |
| Total cholesterol | rs9376090 | C | T | -0.0254 | 0.004 | 2.60E-09 |
| Total cholesterol | rs112201728 | T | C | 0.0581 | 0.0099 | 1.20E-08 |
| Total cholesterol | rs11753995 | A | G | 0.0489 | 0.0048 | 1.84E-23 |
| Total cholesterol | rs3780181 | G | A | -0.0442 | 0.0071 | 6.67E-10 |
| Total cholesterol | rs10900221 | A | G | 0.0255 | 0.0041 | 7.96E-09 |
| Total cholesterol | rs6573778 | C | T | -0.0263 | 0.0039 | 2.96E-11 |
| Total cholesterol | rs10468017 | T | C | 0.0617 | 0.004 | 7.23E-48 |
| Total cholesterol | rs633695 | G | A | 0.0433 | 0.0058 | 1.05E-14 |
| Total cholesterol | rs314253 | C | T | -0.0233 | 0.0037 | 2.81E-10 |
| Total cholesterol | rs7412 | T | C | -0.3736 | 0.0096 | 1.00E-200 |
| Total cholesterol | rs4253772 | T | C | 0.0322 | 0.0058 | 9.85E-09 |
| Total cholesterol | rs515135 | C | T | 0.1238 | 0.0046 | 6.38E-151 |
| Triglycerides | rs1260326 | C | T | -0.1148 | 0.0034 | 1.00E-200 |
| Triglycerides | rs13389219 | T | C | -0.0271 | 0.0034 | 2.60E-15 |
| Triglycerides | rs645040 | T | G | 0.0293 | 0.004 | 1.83E-12 |
| Triglycerides | rs6882076 | C | T | 0.0286 | 0.0035 | 1.51E-15 |
| Triglycerides | rs634869 | C | T | -0.0272 | 0.0033 | 1.78E-14 |
| Triglycerides | rs287621 | C | T | -0.0222 | 0.0037 | 7.67E-09 |
| Triglycerides | rs749671 | A | G | -0.0211 | 0.0034 | 6.11E-10 |
| Triglycerides | rs7248104 | A | G | -0.0222 | 0.0034 | 5.04E-10 |
| Triglycerides | rs3760627 | C | T | 0.0189 | 0.0034 | 5.29E-09 |
| Triglycerides | rs6029143 | T | C | -0.0388 | 0.0071 | 4.93E-08 |
| Triglycerides | rs4810479 | T | C | -0.0474 | 0.0038 | 2.07E-34 |
| Triglycerides | rs12748152 | T | C | 0.0372 | 0.0059 | 1.10E-09 |
| Triglycerides | rs2972146 | T | G | 0.0281 | 0.0034 | 2.97E-15 |
| Triglycerides | rs2239520 | A | G | -0.0236 | 0.0037 | 4.14E-10 |
| Triglycerides | rs998584 | A | C | 0.0293 | 0.0037 | 3.42E-15 |
| Triglycerides | rs4719841 | G | A | 0.0232 | 0.0034 | 8.86E-11 |
| Triglycerides | rs10501321 | C | T | -0.0216 | 0.0035 | 1.41E-08 |
| Triglycerides | rs247616 | T | C | -0.0393 | 0.0037 | 1.12E-25 |
| Triglycerides | rs3761445 | A | G | 0.0232 | 0.0034 | 8.06E-12 |
| Triglycerides | rs17513135 | T | C | 0.0220 | 0.0039 | 1.63E-08 |
| Triglycerides | rs4587594 | A | G | -0.0694 | 0.0035 | 3.50E-82 |
| Triglycerides | rs10440120 | A | C | -0.0306 | 0.0044 | 5.34E-11 |
| Triglycerides | rs6831256 | G | A | 0.0258 | 0.0035 | 1.60E-12 |
| Triglycerides | rs9686661 | T | C | 0.0379 | 0.0044 | 2.54E-16 |
| Triglycerides | rs719726 | T | C | 0.0199 | 0.0035 | 2.49E-08 |
| Triglycerides | rs38855 | G | A | -0.0187 | 0.0033 | 2.11E-08 |
| Triglycerides | rs6995541 | G | A | 0.0265 | 0.0037 | 1.34E-12 |
| Triglycerides | rs12678919 | G | A | -0.1702 | 0.0056 | 1.82E-199 |
| Triglycerides | rs4738684 | G | A | -0.0205 | 0.0035 | 8.82E-09 |
| Triglycerides | rs2954022 | A | C | -0.0780 | 0.0033 | 2.23E-113 |
| Triglycerides | rs1832007 | G | A | -0.0327 | 0.0047 | 1.72E-12 |
| Triglycerides | rs2250802 | A | G | 0.0230 | 0.0037 | 1.21E-10 |
| Triglycerides | rs948690 | C | T | -0.0306 | 0.0052 | 6.57E-09 |
| Triglycerides | rs11613352 | T | C | -0.0280 | 0.0039 | 9.40E-14 |
| Triglycerides | rs2043085 | C | T | -0.0327 | 0.0034 | 7.81E-20 |
| Triglycerides | rs588136 | T | C | -0.0495 | 0.0041 | 3.37E-30 |
| Triglycerides | rs8077889 | C | A | 0.0252 | 0.0042 | 9.88E-09 |
| Triglycerides | rs676210 | A | G | -0.0733 | 0.0039 | 3.28E-71 |
| Triglycerides | rs442177 | T | G | 0.0309 | 0.0033 | 1.32E-18 |
| Triglycerides | rs2665357 | C | A | 0.0212 | 0.0033 | 8.33E-10 |
| Triglycerides | rs11974409 | G | A | -0.0899 | 0.0042 | 1.36E-100 |
| Triglycerides | rs12676857 | C | T | 0.0332 | 0.0046 | 7.29E-12 |
| Triglycerides | rs2068888 | A | G | -0.0241 | 0.0034 | 1.68E-11 |
| Triglycerides | rs11057408 | T | G | -0.0258 | 0.0035 | 2.05E-12 |
| Triglycerides | rs16948098 | A | G | 0.0800 | 0.0089 | 4.84E-17 |
| Triglycerides | rs439401 | C | T | 0.0659 | 0.0038 | 1.42E-66 |
| Triglycerides | rs1321257 | A | G | -0.0402 | 0.0034 | 5.99E-31 |
| Triglycerides | rs2247056 | C | T | 0.0378 | 0.0039 | 3.86E-21 |
| Triglycerides | rs10761762 | C | T | -0.0270 | 0.0033 | 1.06E-17 |
| Triglycerides | rs174535 | C | T | 0.0470 | 0.0034 | 1.73E-41 |
| Triglycerides | rs7350481 | C | T | -0.2254 | 0.0066 | 1.00E-200 |
| Triglycerides | rs12280753 | T | C | 0.1931 | 0.0064 | 1.22E-179 |
| Triglycerides | rs3198697 | T | C | -0.0198 | 0.0034 | 2.21E-08 |
| Triglycerides | rs10401969 | C | T | -0.1210 | 0.0065 | 9.70E-70 |
| Triglycerides | rs731839 | A | G | -0.0224 | 0.0036 | 2.65E-09 |
| Apolipoprotein A | rs1883025 | T | C | -0.0798 | 0.013321 | 2.86E-09 |
| Apolipoprotein A | rs261291 | C | T | 0.1443 | 0.010905 | 2.58E-39 |
| Apolipoprotein A | rs75835816 | C | G | -0.2210 | 0.038801 | 1.67E-08 |
| Apolipoprotein A | rs4860951 | T | A | 0.0737 | 0.013224 | 3.29E-08 |
| Apolipoprotein A | rs144064722 | G | A | 0.2037 | 0.035039 | 8.30E-09 |
| Apolipoprotein A | rs11632618 | A | G | 0.1741 | 0.024085 | 7.76E-13 |
| Apolipoprotein A | rs6507939 | C | A | 0.1084 | 0.014268 | 5.03E-14 |
| Apolipoprotein A | rs1461729 | G | A | 0.0864 | 0.015193 | 1.77E-08 |
| Apolipoprotein A | rs174594 | A | C | 0.0717 | 0.010504 | 1.32E-11 |
| Apolipoprotein A | rs73424577 | G | A | 0.1860 | 0.030161 | 9.78E-10 |
| Apolipoprotein B | rs190934192 | A | G | -0.3197 | 0.040849 | 1.30E-14 |
| Apolipoprotein B | rs11591147 | T | G | -0.4379 | 0.035298 | 2.50E-34 |
| Apolipoprotein B | rs3005923 | A | G | -0.2829 | 0.03695 | 4.73E-14 |
| Apolipoprotein B | rs1367117 | A | G | 0.1089 | 0.011191 | 9.99E-22 |
| Apolipoprotein B | rs635634 | T | C | 0.0740 | 0.012612 | 7.71E-09 |
| Apolipoprotein B | rs7412 | T | C | -0.4276 | 0.025979 | 4.39E-59 |
| Apolipoprotein B | rs1081105 | C | A | 0.2229 | 0.039281 | 2.32E-08 |
| Apolipoprotein B | rs629301 | T | G | 0.0901 | 0.012193 | 3.56E-13 |
| Apolipoprotein B | rs182695896 | C | A | 0.2365 | 0.041781 | 2.49E-08 |
| Apolipoprotein B | rs142130958 | A | G | -0.1996 | 0.016743 | 7.78E-32 |
| Apolipoprotein B | rs2495477 | G | A | -0.0619 | 0.011013 | 3.14E-08 |
| Apolipoprotein B | rs1260326 | C | T | -0.0668 | 0.010393 | 2.51E-10 |
| Apolipoprotein B | rs6756629 | A | G | -0.1133 | 0.018566 | 1.90E-09 |
| Apolipoprotein B | rs4722043* | C | G | -0.0659 | 0.010669 | 1.17E-09 |
| Apolipoprotein B | rs115849089 | A | G | -0.0996 | 0.017126 | 1.04E-08 |
| Apolipoprotein B | rs10056811 | A | G | 0.0858 | 0.010571 | 1.35E-15 |
| Apolipoprotein B | rs150617279* | A | T | -0.1124 | 0.017688 | 3.97E-10 |
| Apolipoprotein B | rs144064722 | G | A | 0.1990 | 0.035059 | 2.29E-08 |
| Apolipoprotein B | rs2980875 | G | A | -0.0697 | 0.009993 | 6.68E-12 |
| Apolipoprotein B | rs964184* | C | G | -0.1658 | 0.014266 | 2.58E-30 |
| Apolipoprotein B | rs1883711* | C | G | 0.1441 | 0.02526 | 1.95E-08 |

EA= effect allele or low-density lipoprotein-cholesterol (LDL-C) lowering allele; OA = other or non-effect allele; EAF = effect allele frequency; se= standard error. * Palindromic SNPs removed.

Beta represents the change in lipid trait levels per copy of the effect allele.

**S2.1-2.5 Tables.** Genetic correlation results for HMGCR, NPC1L1, CETP, PCSK9 and LDLR single nucleotide polymorphisms.

**S2.1 Table.**

| ***HMGCR*** | rs10066707 | rs2303152 | rs17238484 | rs5909 | rs12916 |
| --- | --- | --- | --- | --- | --- |
| rs10066707 | 1 | 0.068 | 0.266 | 0.106 | 0.31 |
| rs2303152 | 0.068 | 1 | 0.317 | 0.013 | 0.13 |
| rs17238484 | 0.266 | 0.317 | 1 | 0.04 | 0.378 |
| rs5909 | 0.106 | 0.013 | 0.04 | 1 | 0.213 |
| rs12916 | 0.31 | 0.13 | 0.378 | 0.213 | 1 |

**S2.2 Table.**

| ***NPC1L1*** | rs10234070 | rs2073547 | rs217386 | rs7791240 | rs2300414 |
| --- | --- | --- | --- | --- | --- |
| rs10234070 | 1 | 0.198 | 0.048 | 0 | 0.002 |
| rs2073547 | 0.198 | 1 | 0.135 | 0.295 | 0.08 |
| rs217386 | 0.048 | 0.135 | 1 | 0.077 | 0.042 |
| rs7791240 | 0 | 0.295 | 0.077 | 1 | 0.36 |
| rs2300414 | 0.002 | 0.08 | 0.042 | 0.36 | 1 |

**S2.3 Table.**

| **CETP** | **rs9989419** | **rs12708967** | **rs3764261** | **rs1800775** | **rs1864163** | **rs289714** |
| --- | --- | --- | --- | --- | --- | --- |
| **rs9989419** | 1 | 0.124 | 0.232 | 0.272 | 0.259 | 0.089 |
| **rs12708967** | 0.124 | 1 | 0.105 | 0.033 | 0.191 | 0.198 |
| **rs3764261** | 0.232 | 0.105 | 1 | 0.469 | 0.179 | 0.045 |
| **rs1800775** | 0.272 | 0.033 | 0.469 | 1 | 0.381 | 0.135 |
| **rs1864163** | 0.259 | 0.191 | 0.179 | 0.381 | 1 | 0.418 |
| **rs289714** | 0.089 | 0.198 | 0.045 | 0.135 | 0.418 | 1 |

**S2.4 Table.**

| ***PCSK9*** | rs2479394 | rs11206510 | rs2479409 | rs10888897 | rs7552841 | rs562556 |
| --- | --- | --- | --- | --- | --- | --- |
| rs2479394 | 1 | 0.053 | 0.011 | 0.002 | 0.03 | 0.006 |
| rs11206510 | 0.053 | 1 | 0.067 | 0.068 | 0.018 | 0 |
| rs2479409 | 0.011 | 0.067 | 1 | 0.09 | 0 | 0.001 |
| rs10888897 | 0.002 | 0.068 | 0.09 | 1 | 0 | 0.047 |
| rs7552841 | 0.03 | 0.018 | 0 | 0 | 1 | 0.063 |
| rs562556 | 0.006 | 0 | 0.001 | 0.047 | 0.063 | 1 |

**S2.5 Table.**

| ***LDLR*** | rs1122608 | rs6511720 | rs688 |
| --- | --- | --- | --- |
| rs1122608 | 1 | 0.202 | 0.084 |
| rs6511720 | 0.202 | 1 | 0.006 |
| rs688 | 0.084 | 0.006 | 1 |

**S3 Table.** Mendelian randomization results of genetically-proxied inhibition of *HMGCR,* *NPC1L1, CETP, PCSK9* and *LDLR* with risk of combined oral/ oropharyngeal cancer accounting for LD structure in GAME-ON.

| **Outcome** | **Exposure** | **Method** | **Beta** | **se** | **OR** | **CIL** | **CIU** | **P-value** |
| --- | --- | --- | --- | --- | --- | --- | --- | --- |
| OC and OPC | *HMGCR* | IVW | 0.0529 | 0.370 | 1.05 | 0.51 | 2.18 | 0.89 |
| OC and OPC | *HMGCR* | MR Egger | 0.5254 | 1.344 | 1.69 | 0.12 | 23.54 | 0.70 |
| OC and OPC | *HMGCR* | Weighted median | 0.1797 | 0.331 | 1.20 | 0.63 | 2.29 | 0.59 |
| OC and OPC | *NPC1L1* | IVW | -0.0068 | 0.595 | 0.99 | 0.31 | 3.19 | 0.99 |
| OC and OPC | *NPC1L1* | MR Egger | -1.5396 | 6.262 | 0.21 | 1.00E-06 | 4.59E+04 | 0.81 |
| OC and OPC | *NPC1L1* | Weighted median | -0.1059 | 0.583 | 0.90 | 0.29 | 2.82 | 0.86 |
| OC and OPC | *CETP* | IVW | 0.3201 | 0.4943 | 1.38 | 0.52 | 3.63 | 0.52 |
| OC and OPC | *CETP* | MR Egger | 0.2994 | 1.8661 | 1.35 | 0.03 | 52.30 | 0.87 |
| OC and OPC | *CETP* | Weighted median | 0.2133 | 0.4271 | 1.24 | 0.54 | 2.86 | 0.62 |
| OC and OPC | *PCSK9* | IVW | 0.7086 | 0.275 | 2.03 | 1.18 | 3.48 | 0.01 |
| OC and OPC | *PCSK9* | MR Egger | 0.5135 | 1.137 | 1.67 | 0.18 | 15.53 | 0.65 |
| OC and OPC | *PCSK9* | Weighted median | 0.7934 | 0.318 | 2.21 | 1.18 | 4.13 | 0.01 |
| OC and OPC | *LDLR* | IVW | -0.3696 | 0.216 | 0.69 | 0.45 | 1.06 | 0.09 |
| OC and OPC | *LDLR* | MR Egger | -0.0518 | 0.372 | 0.95 | 0.46 | 1.97 | 0.89 |
| OC and OPC | *LDLR* | Weighted median | -0.3417 | 0.216 | 0.71 | 0.47 | 1.09 | 0.11 |

Abbreviations: se, standard error; OR, odds ratio; CIL, lower confidence interval; CIU, upper confidence interval

**S4 Table.** Assessing weak instrument bias (F-statistic) and proportion of variance in the phenotype (*R*^2^) explained by the genetic instruments.

|  | R^2^ | F-statistic |
| --- | --- | --- |
| ***HMGCR*** | 0.003 | 111 |
| ***NPC1L1*** | 0.001 | 51 |
| ***CETP*** | 0.003 | 68 |
| ***PCSK9*** | 0.005 | 140 |
| ***LDLR*** | 0.008 | 508 |
| **HDL-C** | 0.05 | 108 |
| **LDL-C** | 0.06 | 140 |
| **Total cholesterol** | 0.06 | 132 |
| **Total triglycerides** | 0.04 | 163 |
| **Apolipoprotein A** | 0.004 | 83 |
| **Apolipoprotein B** | 0.007 | 72 |

**S5 Table.** Assessing heterogeneity of single nucleotide polymorphism effect estimates in inverse-variance weighted (IVW) and MR Egger regression for primary analysis.

| **Exposure** | **Exposure dataset** | **Q IVW** | **df** | **P** | **Q MR Egger** | **df** | **P** |
| --- | --- | --- | --- | --- | --- | --- | --- |
| *HMGCR* | GLGC[25] | 2.75 | 4 | 0.60 | 2.73 | 3 | 0.44 |
| *NPC1L1* | GLGC[25] | 0.76 | 4 | 0.94 | 0.52 | 3 | 0.92 |
| *CETP* | GLGC[25] | 1.99 | 5 | 0.85 | 1.92 | 4 | 0.75 |
| *PCSK9* | GLGC[25] | 4.61 | 5 | 0.47 | 4.60 | 4 | 0.33 |
| *LDLR* | GLGC[25] | 2.33 | 2 | 0.31 | 1.18 | 1 | 0.28 |

Abbreviations: Q, Q-statistic; df, degrees of freedom; P, p-value.

**S6 Table.** Assessing heterogeneity of single nucleotide polymorphism effect estimates in inverse-variance weighted (IVW) and MR Egger regression for secondary analysis .

| **Exposure** | **Exposure dataset** | **Q IVW** | **df** | **P** | **Q MR Egger** | **df** | **P** |
| --- | --- | --- | --- | --- | --- | --- | --- |
| HDL-C | GLGC[25] | 115.65 | 84 | 0.01 | 115.56 | 83 | 0.01 |
| LDL-C | GLGC[25] | 86.91 | 76 | 0.18 | 86.87 | 75 | 0.16 |
| Total cholesterol | GLGC[25] | 92.58 | 81 | 0.18 | 92.22 | 80 | 0.17 |
| Total triglycerides | GLGC[25] | 59.01 | 53 | 0.27 | 58.41 | 52 | 0.25 |
| Apolipoprotein A | 14 studies (Kettunen et al.)[33] | 7.39 | 8 | 0.50 | 6.65 | 7 | 0.47 |
| Apolipoprotein B | 14 studies (Kettunen et al.)[33] | 18.11 | 13 | 0.15 | 18.05 | 12 | 0.11 |

Abbreviations: Q, Q-statistic; df, degrees of freedom; P, p-value.

**Supplementary Table 7** Assessing directional pleiotropy through MR Egger intercept for primary analysis

| **Exposure** | **Exposure dataset** | **N SNPs** | **Estimate** | **SE** | **P** |
| --- | --- | --- | --- | --- | --- |
| *HMGCR* | GLGC[25] | 5 | 0.011 | 0.076 | 0.89 |
| *NPC1L1* | GLGC[25] | 5 | -0.049 | 0.099 | 0.65 |
| *CETP* | GLGC^22^ | 6 | -0.016 | 0.062 | 0.81 |
| *PCSK9* | GLGC[25] | 6 | -0.006 | 0.059 | 0.93 |
| *LDLR* | GLGC[25] | 3 | 0.035 | 0.035 | 0.51 |

Abbreviations: UVMR, univariable Mendelian randomization; SE, standard error; P, p-value.

**Supplementary Table 8** Assessing directional pleiotropy through MR Egger intercept for secondary analysis

| **Exposure** | **Exposure dataset** | **N SNPs** | **Estimate** | **SE** | **P** |
| --- | --- | --- | --- | --- | --- |
| LDL-C | GLGC[25] | 77 | 0.001 | 0.006 | 0.86 |
| HDL-C | GLGC[25] | 85 | -0.002 | 0.007 | 0.80 |
| Total cholesterol | GLGC[25] | 82 | -0.003 | 0.006 | 0.58 |
| Total triglycerides | GLGC[25] | 54 | -0.005 | 0.007 | 0.47 |
| Apolipoprotein A | 14 studies (Kettunen et al.)[33] | 9 | 0.029 | 0.033 | 0.42 |
| Apolipoprotein B | 14 studies (Kettunen et al.)[33] | 14 | -0.004 | 0.019 | 0.85 |

Abbreviations: UVMR, univariable Mendelian randomization; SE, standard error; P, p-value.

**S9 Table.** MR-PRESSO results for HMGCR, NPC1L1, CETP, PCSK9, LDLR and other lipid trait SNPs on combined oral/ oropharyngeal cancer.

| **Outcome** | **Exposure** | **RSSobs** | **P-value** |
| --- | --- | --- | --- |
| OC and OPC | *HMGCR* | 3.67 | 6.91E-01 |
| OC and OPC | *NPC1L1* | 1.06 | 9.58E-01 |
| OC and OPC | *CETP* | 2.85 | 8.44 E-01 |
| OC and OPC | *PCSK9* | 6.52 | 4.84E-01 |
| OC and OPC | *LDLR* | NA (insufficient SNPs) | NA (insufficient SNPs) |
| OC and OPC | LDL-C | 90.49 | 0.16 |
| OC and OPC | HDL-C | 117.57 | 0.01 |
| OC and OPC | Total cholesterol | 95.11 | 0.181 |
| OC and OPC | Total triglycerides | 61.08 | 0.28 |
| OC and OPC | Apolipoprotein A | 8.57 | 0.56 |
| OC and OPC | Apolipoprotein B | 21.82 | 0.13 |

Abbreviations: RSSobs, residual sum of squares observations.

**S10 Table.** Assessing violation of the “NO Measurement Error” (NOME) assumption for instruments used in MR-Egger regression.

| **Exposure** | **Exposure dataset** | **I^2^ unweighted** | **I^2^ weighted** |
| --- | --- | --- | --- |
| *HMGCR* | GLGC[25] | 0.78 | 0.93 |
| *NPC1L1* | GLGC[25] | 0.46 | 0.86 |
| *CETP* | GLGC^22^ | 0.67 | 0.86 |
| *PCSK9* | GLGC[25] | 0.99 | 0.98 |
| *LDLR* | GLGC[25] | 0.99 | 0.99 |
| LDL-C | GLGC[25] | 0.99 | 0.98 |
| HDL-C | GLGC[25] | 0.97 | 0.97 |
| Total cholesterol | GLGC[25] | 0.98 | 0.98 |
| Total triglycerides | GLGC[25] | 0.99 | 0.98 |
| Apolipoprotein A | 14 studies (Kettunen et al.)[33] | 0.92 | 0.95 |
| Apolipoprotein B | 14 studies (Kettunen et al.)[33] | 0.95 | 0.93 |

Abbreviations: I^2^, I-squared statistic.

**S11 Table.** SIMEX correction MR Egger regression results for HMGCR, NPC1L1 and CETP (where I^2^ <0.90).

| **Outcome** | **Exposure** | **OR** | **CIL** | **CIU** | **P** |
| --- | --- | --- | --- | --- | --- |
| OC and OPC | *HMGCR* | 0.60 | 0.03 | 12.20 | 0.76 |
| OC and OPC | *NPC1L1* | 7.77 | 0.19 | 321.65 | 0.36 |
| OC and OPC | *CETP* | 1.82 | 0.03 | 120.28 | 0.79 |

Abbreviations: OR, odds ratio; CIL, lower confidence interval; CIU, upper confidence interval; P, p-value.

**S12 Table.** Mendelian randomization results of genetically-proxied inhibition of HMGCR, NPC1L1, CETP, PCSK9 and LDLR with risk of oral and oropharyngeal cancer including sensitivity analyses in UK Biobank.

|  |  | | | | **IVW** | | **Weighted median** | | **Weighted mode** | | | **MR-Egger** | | |
| --- | --- | --- | --- | --- | --- | --- | --- | --- | --- | --- | --- | --- | --- | --- |
|  | **Outcome** | **Exposure/**  **Outcome**  **dataset** | **Outcome N** | **Number of SNPs** | **OR (95%CI)** | **P** | **OR (95%CI)** | **P** | | **OR (95%CI)** | **P** | | **OR (95%CI)** | **P** |
| *HMGCR* | Oral/ Oropharyngeal cancer | UK Biobank/ GLGC | 839 | 5 | 0.55 (0.21, 1.46) | 0.23 | 0.59 (0.18, 1.89) | 0.38 | | 0.57 (0.14, 2.31) | 0.46 | | 3.76 (0.01, 1145.91) | 0.67 |
|  | Oral cancer | UK Biobank/ GLGC | 357 | 5 | 0.24 (0.05, 1.09) | 0.06 | 0.48 (0.08, 3.03) | 0.44 | | 0.49 (0.05, 4.33) | 0.55 | | 31.78 (0.00, 203203.76) | 0.48 |
|  | Oropharyngeal cancer | UK Biobank/ GLGC | 494 | 5 | 1.01 (0.28, 3.63) | 0.99 | 0.74 (0.15, 3.67) | 0.71 | | 0.75 (0.13, 4.33) | 0.76 | | 0.52 (0.00, 890.93) | 0.87 |
| *NPC1L1* | Oral/ Oropharyngeal cancer | UK Biobank/ GLGC | 839 | 5 | 1.14 (0.05, 24.82) | 0.94 | 2.54 (0.14, 45.32) | 0.52 | | 3.79 (0.05, 284.57) | 0.58 | | 0.02 (0.00, 46457078.93) | 0.76 |
|  | Oral cancer | UK Biobank/ GLGC | 357 | 5 | 2.91 (0.03, 277.95) | 0.65 | 6.99 (0.09, 554.04) | 0.38 | | 22.20 (0.02, 27423.14) | 0.44 | | 0.00 (0.00, 67338766.60) | 0.51 |
|  | Oropharyngeal cancer | UK Biobank/ GLGC | 494 | 5 | 0.50 (0.04, 5.88) | 0.58 | 1.58 (0.08, 32.81) | 0.77 | | 2.72 (0.10, 74.56) | 0.59 | | 4.39 (0.00, 39822777.00) | 0.87 |
| *CETP* | Oral/ Oropharyngeal cancer | UK Biobank/ GLGC | 839 | 5 | 2.66 (0.83, 8.59) | 0.10 | 2.19 (0.49, 9.71) | 0.30 | | 1.74 (0.24, 12.48) | 0.60 | | 9.07 (0.02, 4195.65) | 0.51 |
|  | Oral cancer | UK Biobank/ GLGC | 357 | 6 | 6.84 (1.14, 41.13) | 0.04 | 11.21 (1.16, 108.63) | 0.04 | | 15.89 (0.76, 330.89) | 0.12 | | 5.93 (0.00, 71920.27) | 0.72 |
|  | Oropharyngeal cancer | UK Biobank/ GLGC | 494 | 6 | 0.99 (0.21, 4.54) | 0.99 | 1.29 (0.19, 8.76) | 0.80 | | 1.49 (0.14, 15.60) | 0.75 | | 5.31 (0.00, 15730.94) | 0.70 |
| *PCSK9* | Oral/ Oropharyngeal cancer | UK Biobank/ GLGC | 839 | 5 | 1.17 (0.50, 2.69) | 0.72 | 1.04 (0.37, 2.97) | 0.94 | | 1.05 (0.24, 4.57) | 0.95 | | 0.15 (0.00, 8.67) | 0.40 |
|  | Oral cancer | UK Biobank/ GLGC | 357 | 6 | 0.70 (0.19, 2.53) | 0.59 | 0.53 (0.09, 2.94) | 0.46 | | 0.24 (0.01, 4.20) | 0.36 | | 0.75 (0.00, 374.07) | 0.93 |
|  | Oropharyngeal cancer | UK Biobank/ GLGC | 494 | 6 | 1.43 (0.48, 4.26) | 0.52 | 1.33 (0.34, 5.19) | 0.68 | | 0.80 (0.11, 5.81) | 0.83 | | 0.07 (0.00, 13.21) | 0.36 |
| *LDLR* | Oral/ Oropharyngeal cancer | UK Biobank/ GLGC | 839 | 5 | 0.91 (0.44, 1.90) | 0.80 | 0.93 (0.43, 2.00) | 0.86 | | 0.94 (0.41, 2.16) | 0.91 | | 1.01 (0.27, 3.73) | 0.99 |
|  | Oral cancer | UK Biobank/ GLGC | 357 | 3 | 1.55 (0.50, 4.78) | 0.44 | 1.67 (0.51, 5.43) | 0.40 | | 2.04 (0.53, 7.82) | 0.41 | | 4.03 (0.55, 29.83) | 0.40 |
|  | Oropharyngeal cancer | UK Biobank/ GLGC | 494 | 3 | 0.54 (0.21, 1.39) | 0.20 | 0.47 (0.17, 1.31) | 0.15 | | 0.47 (0.16, 1.37) | 0.30 | | 0.35 (0.06, 1.92) | 0.44 |

Abbreviations: IVW, inverse variance weighted; OR, odds ratio; CI, confidence intervals; P, *p*-value.

OR represents the exponential change in odds of oral/ oropharyngeal squamous cell carcinoma per genetically-proxied inhibition of drug target equivalent to a 1 mmol/L decrease in LDL-C.

**Supplementary Figures**

**S1 Fig.** Forest plots showing Mendelian randomization results for genetically-proxied inhibition of *HMGCR*, *NPC1L1*, *CETP*, *PCSK9* and *LDLR* with risk of combined oral/ oropharyngeal cancer in GAME-ON.

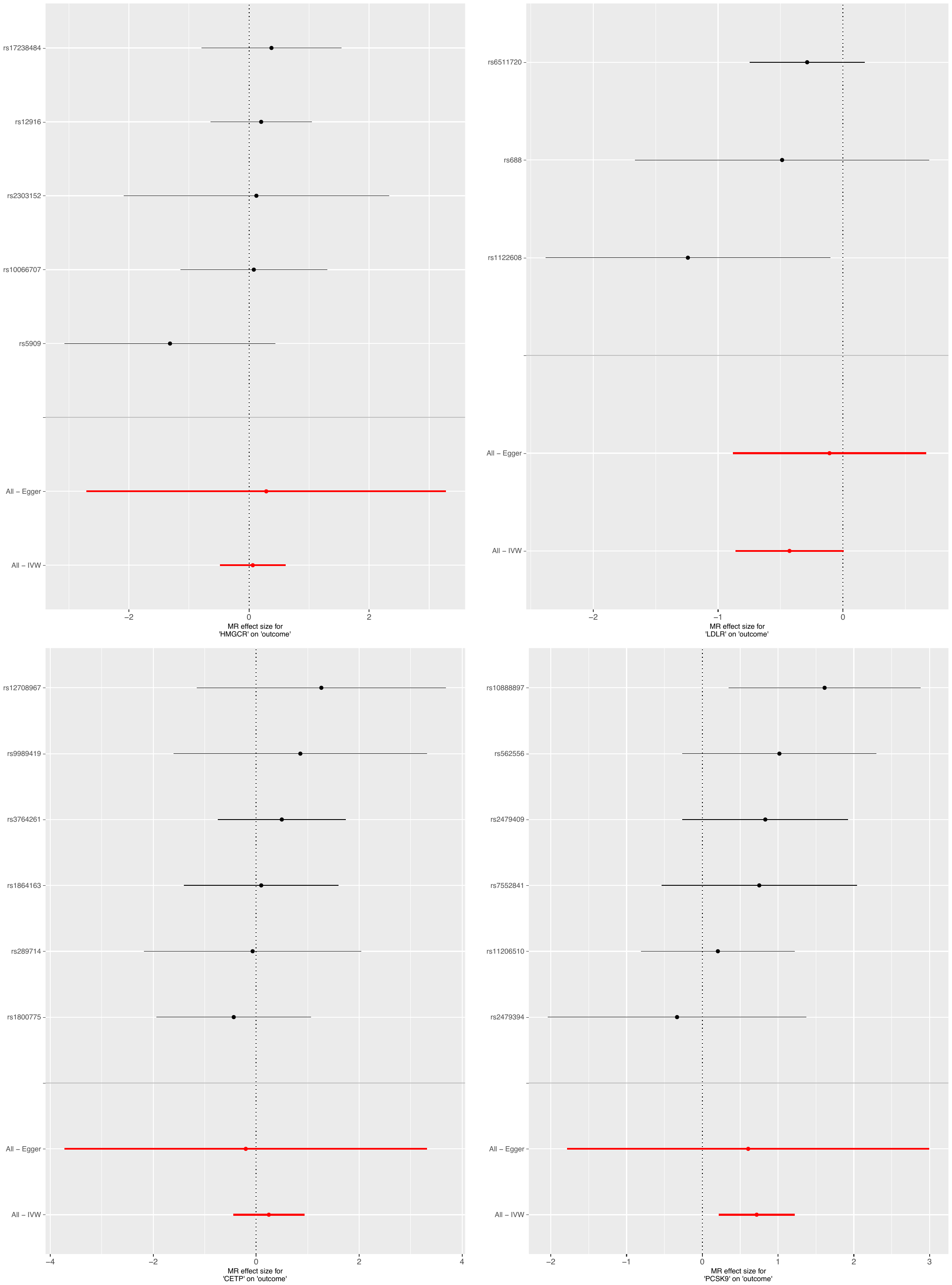

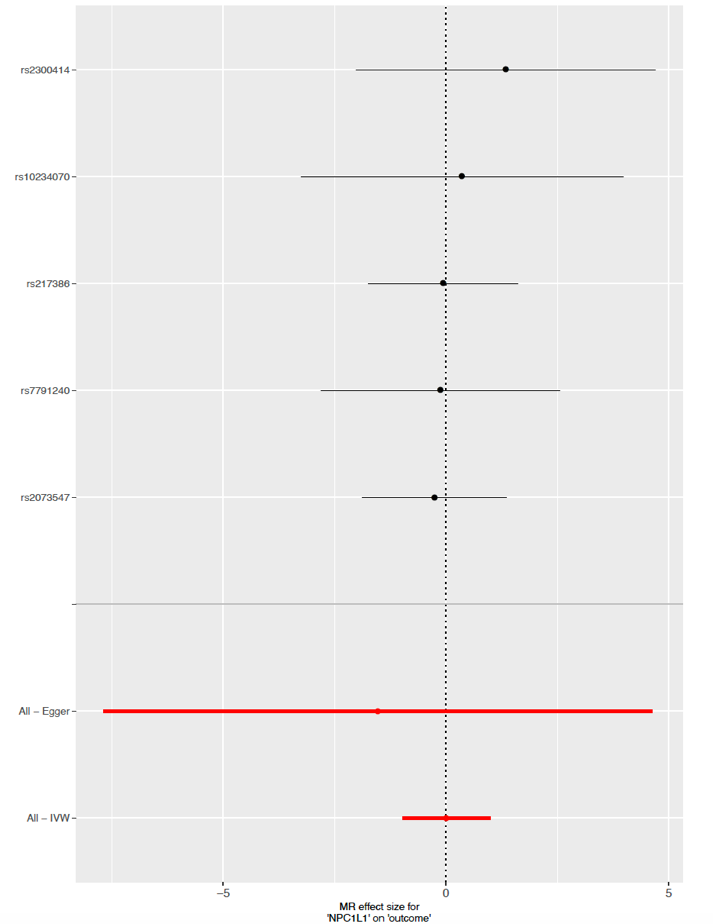

**S2 Fig.** Forest plots showing Mendelian randomization results for LDL-C, HDL-C, total cholesterol, total triglycerides, Apolipoprotein A and Apoprotein B single nucleotide polymorphisms effect on combined oral/oropharyngeal cancer in GAME-ON.

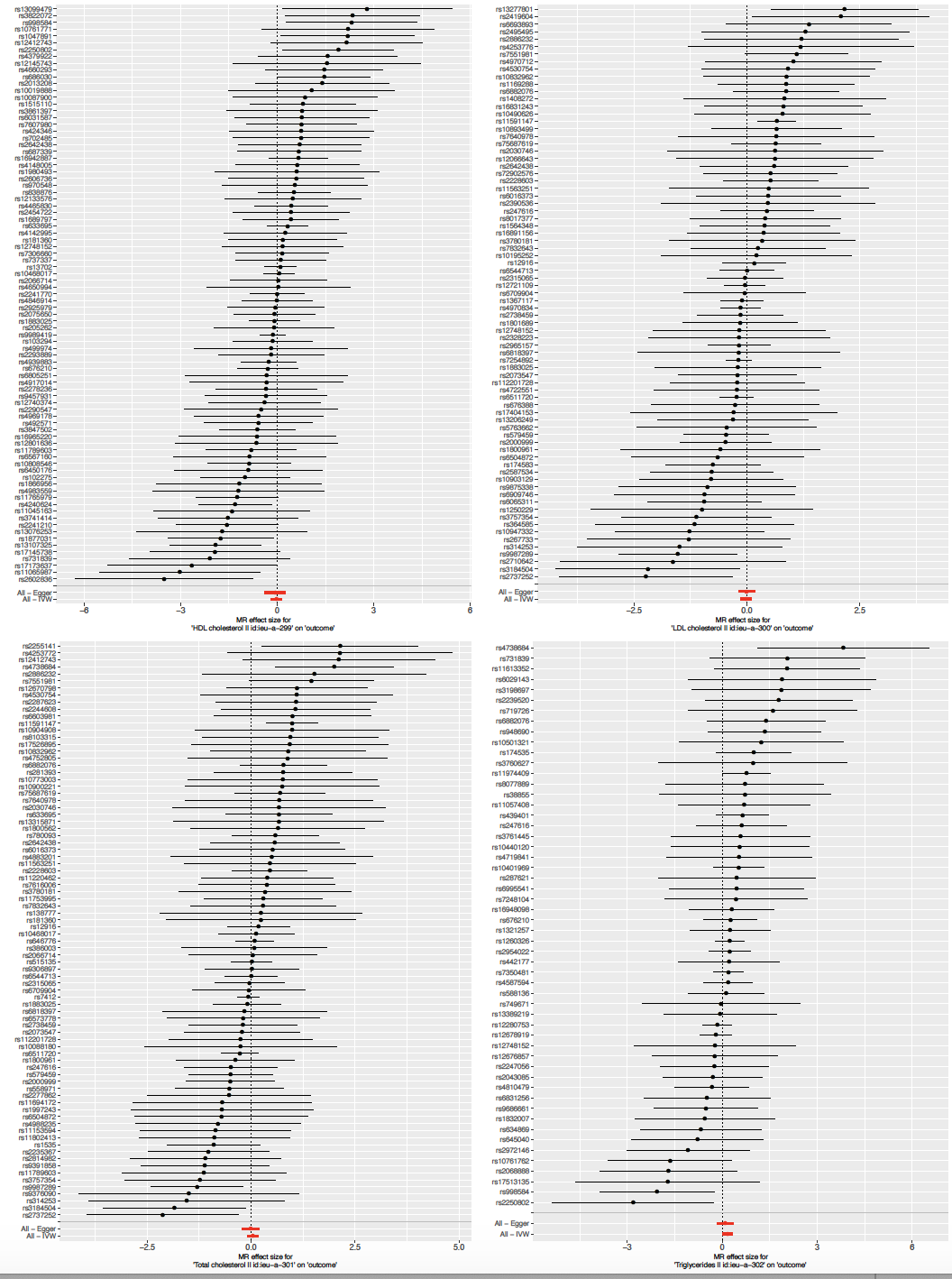

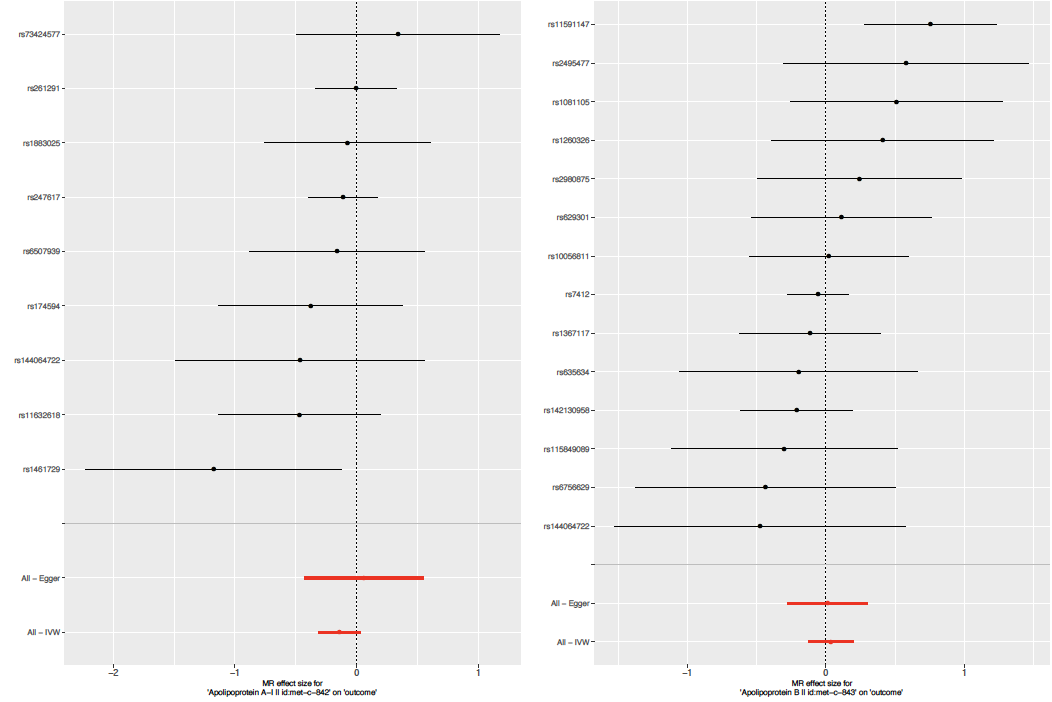

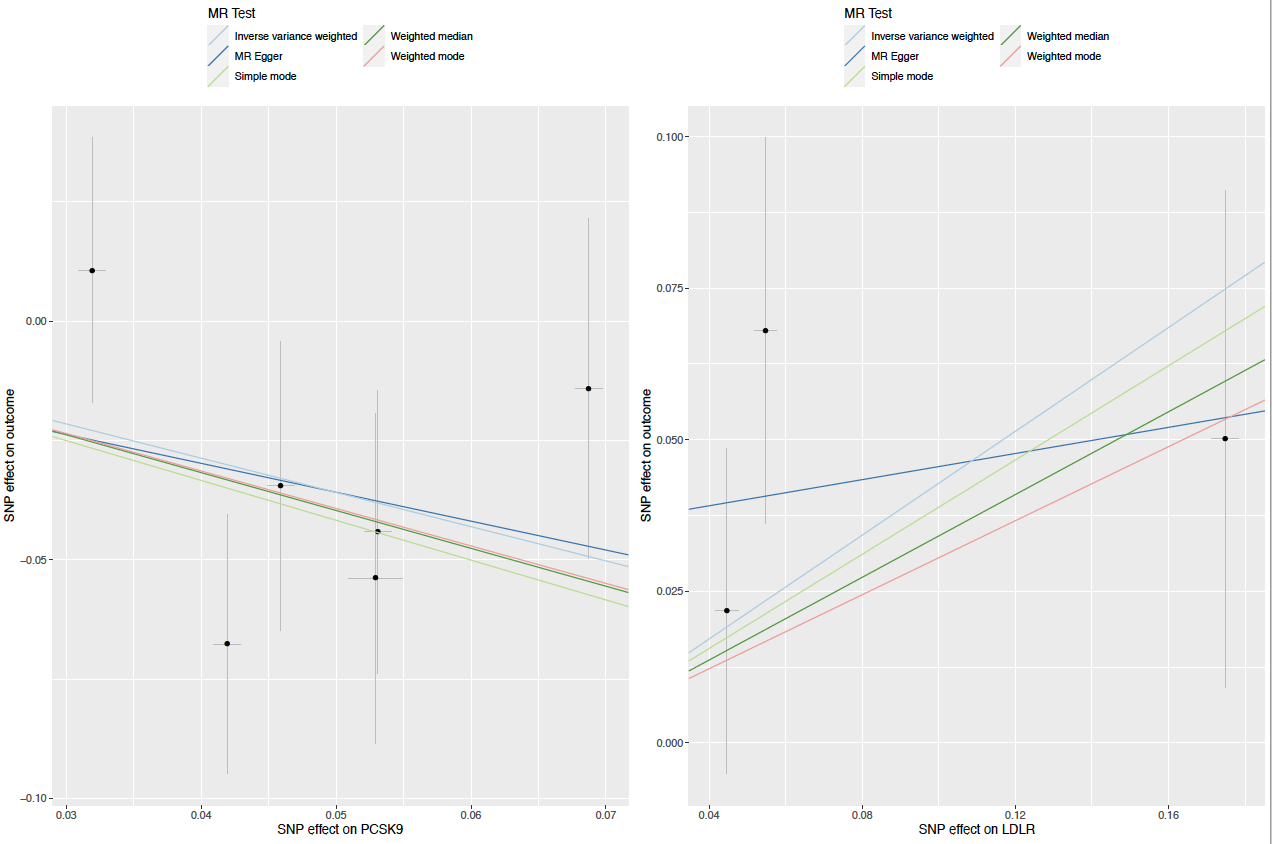
**S3 Fig.** Scatter plots for *HMGCR*, *NPC1L1* and *CETP* single nucleotide polymorphisms effect on combined oral/oropharyngeal cancer in GAME-ON.

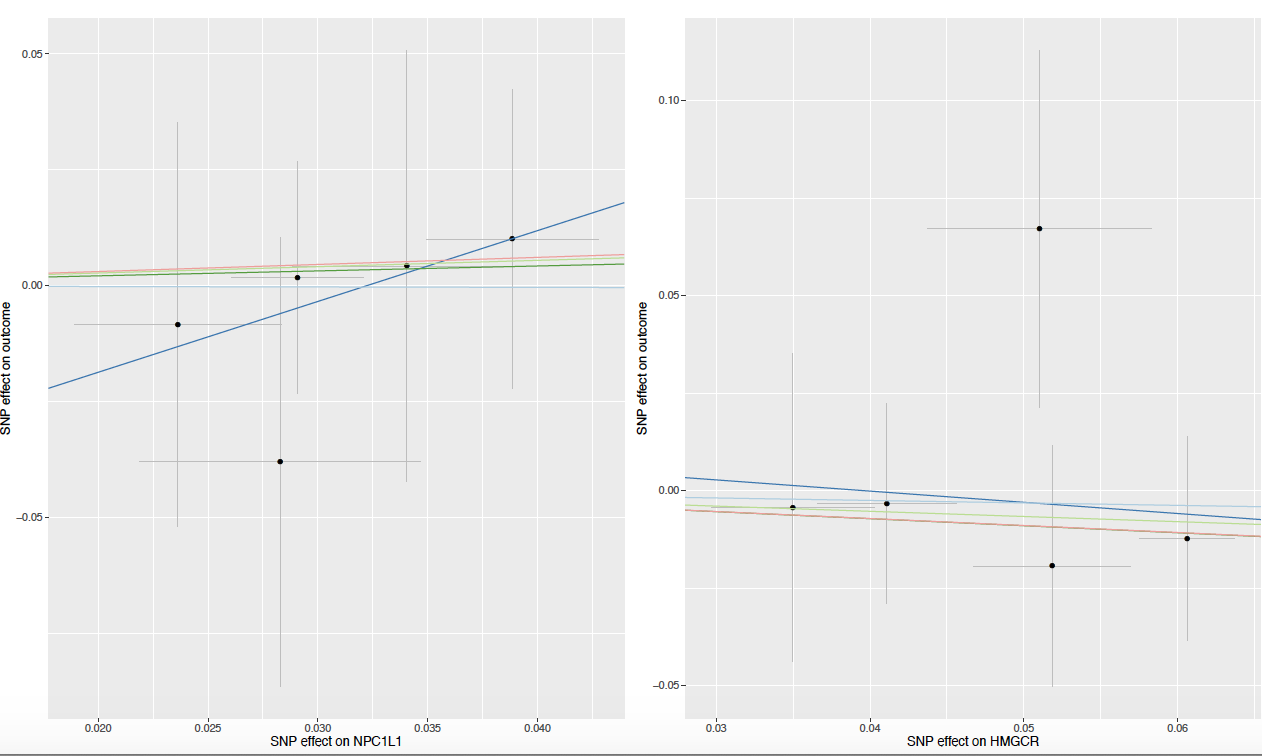

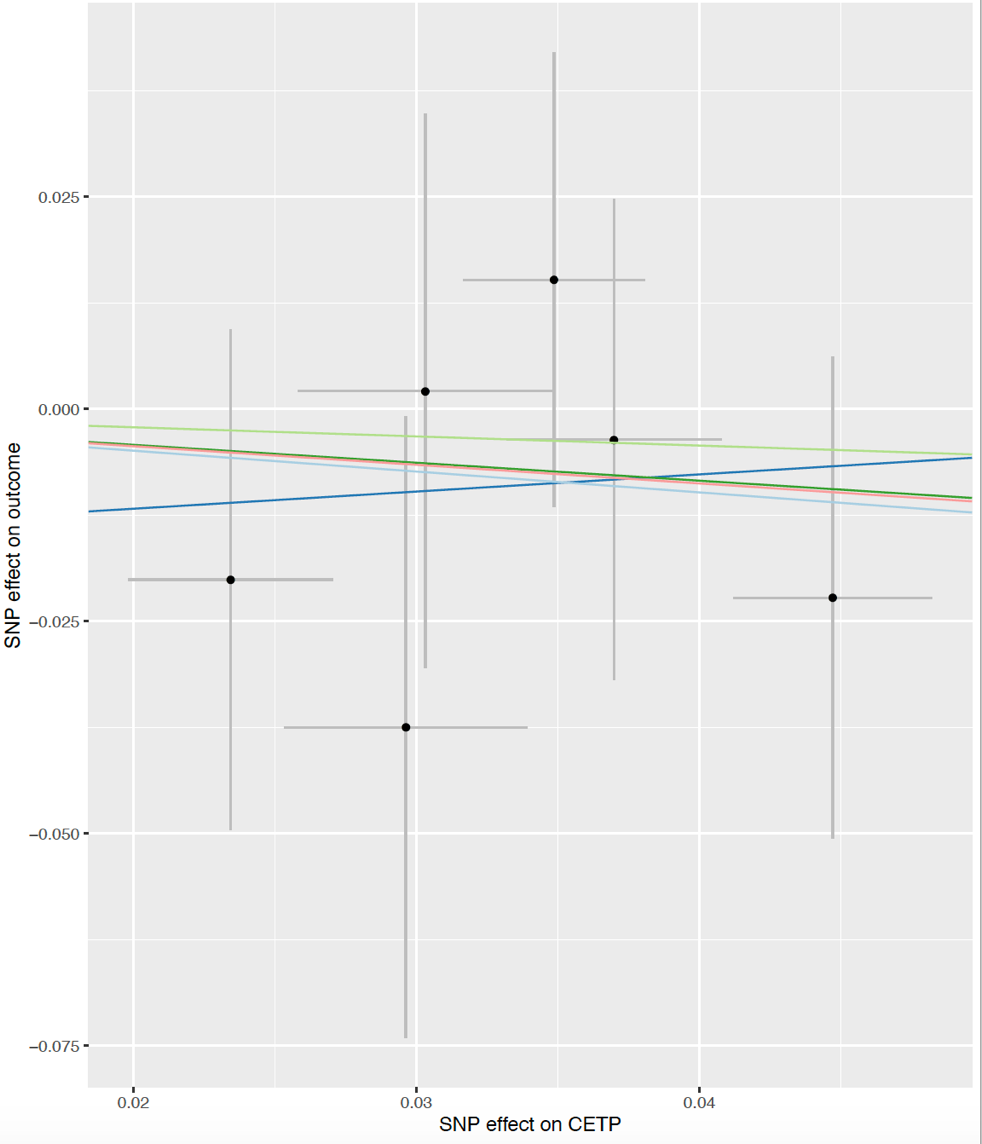

**S4 Fig.** Leave one out analysis for *HMGCR*, *NPC1L1*, *CETP*, *PCSK9* and *LDLR* single nucleotide polymorphisms effect on combined oral/ oropharyngeal cancer in GAME-ON.

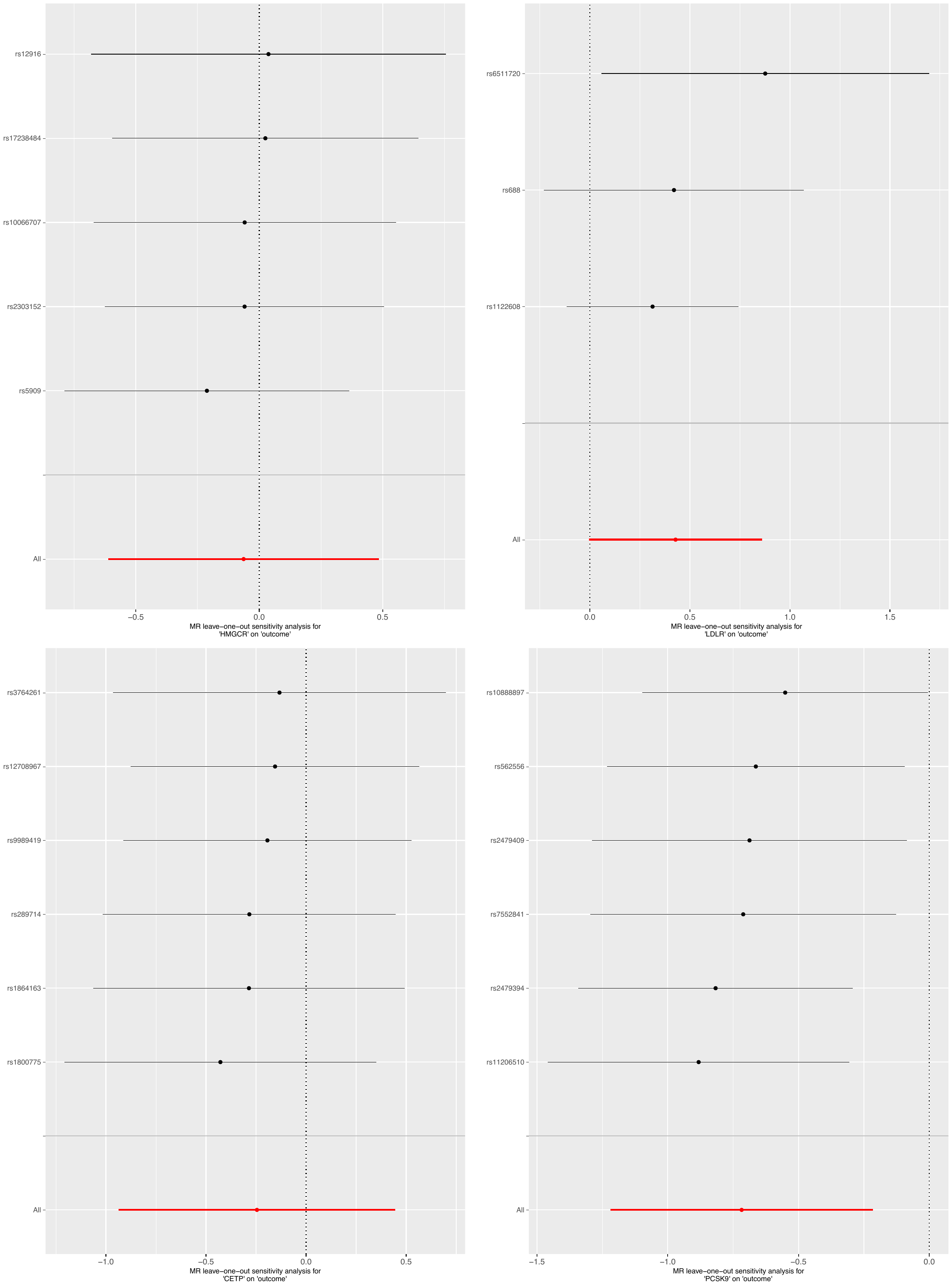

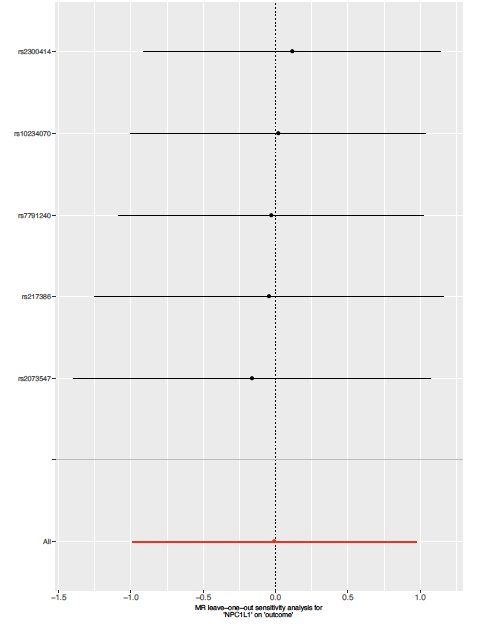

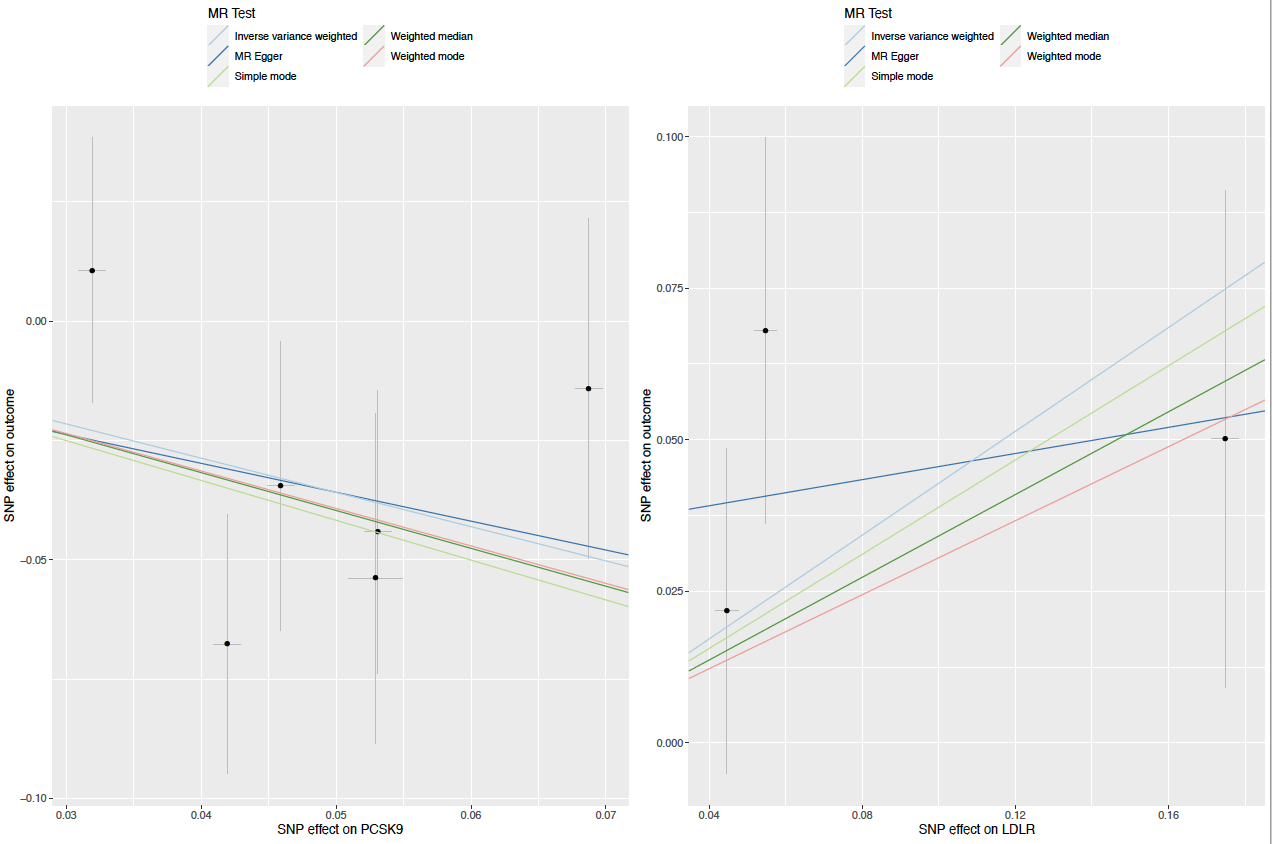
**S5 Fig.** Scatter plots for LDL-C, HDL-C, total cholesterol, total triglycerides, apolipoprotein A and apolipoprotein B single nucleotide polymorphisms effect on combined oral/oropharyngeal cancer in GAME-ON.

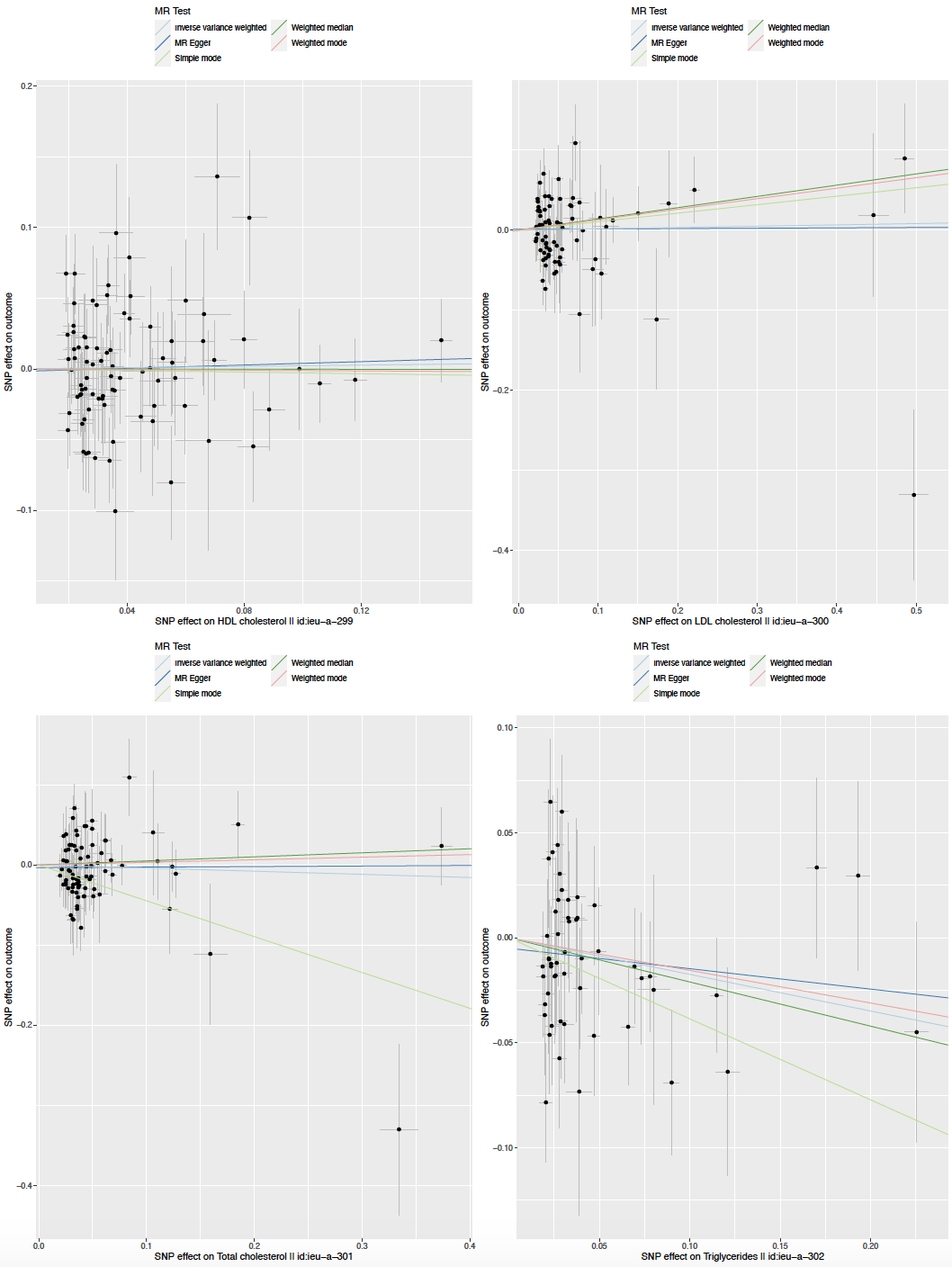

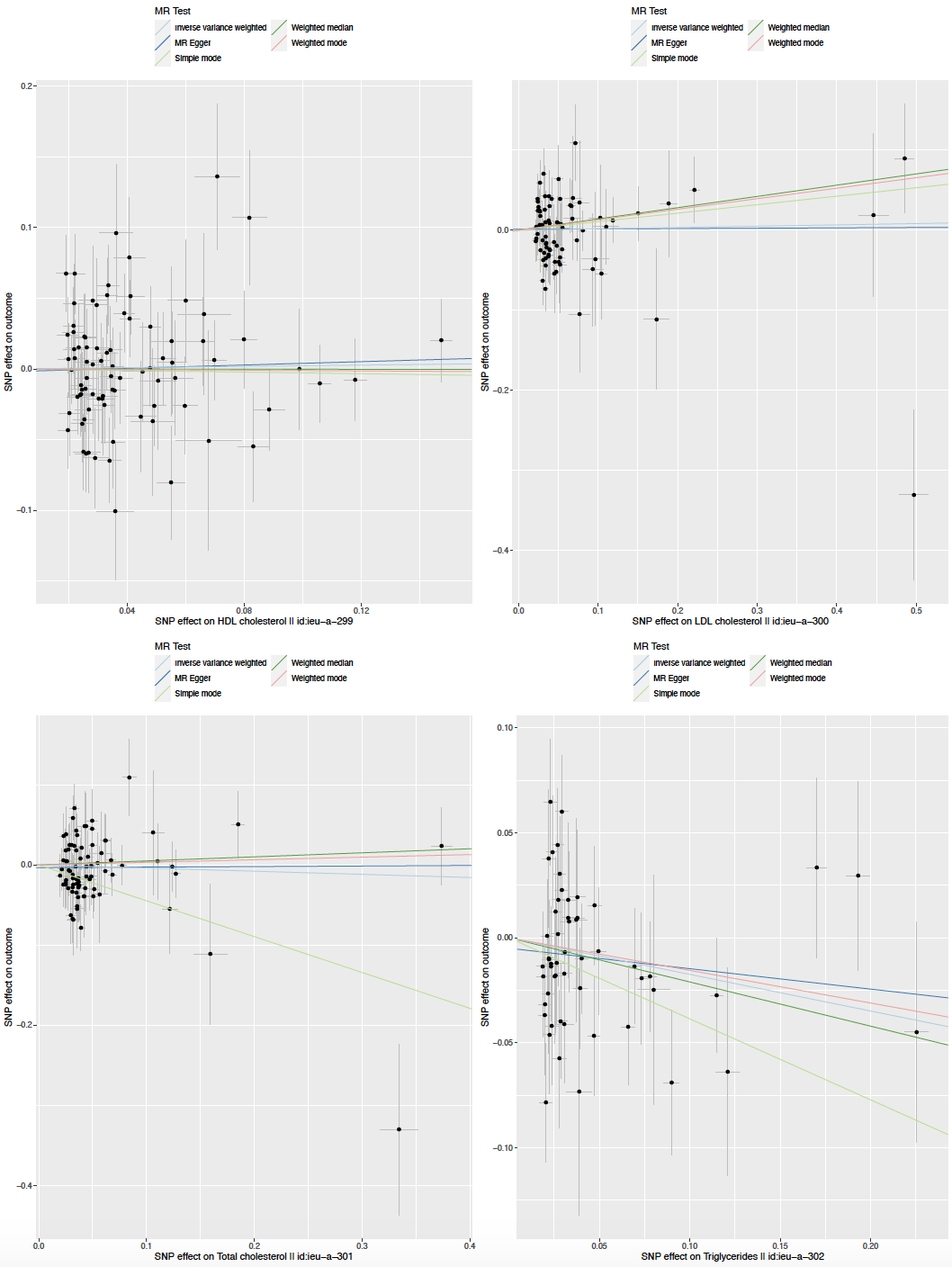

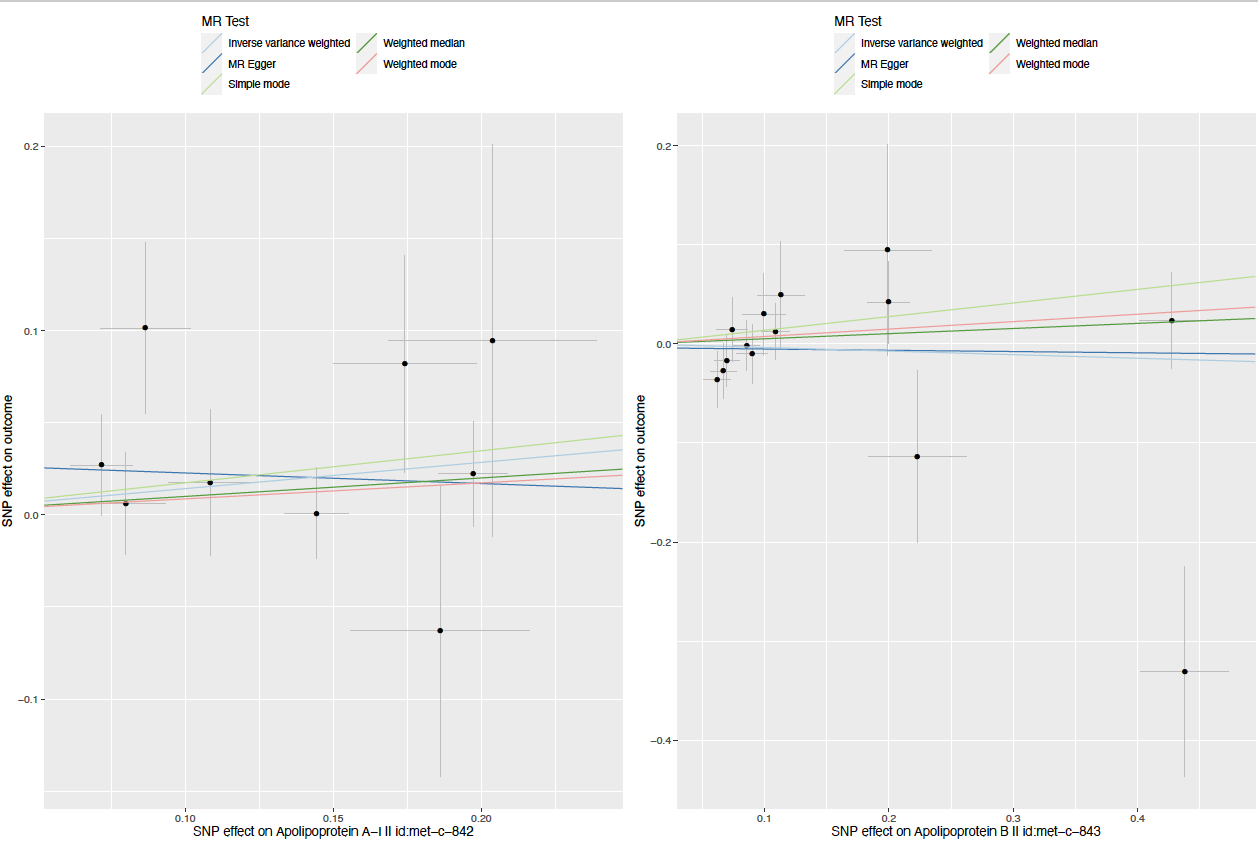

**S6 Fig.** Leave one out analysis for LDL-C, HDL-C, total cholesterol, total triglycerides, Apolipoprotein A and Apoprotein B single nucleotide polymorphisms effect on combined oral/oropharyngeal cancer in GAME-ON.

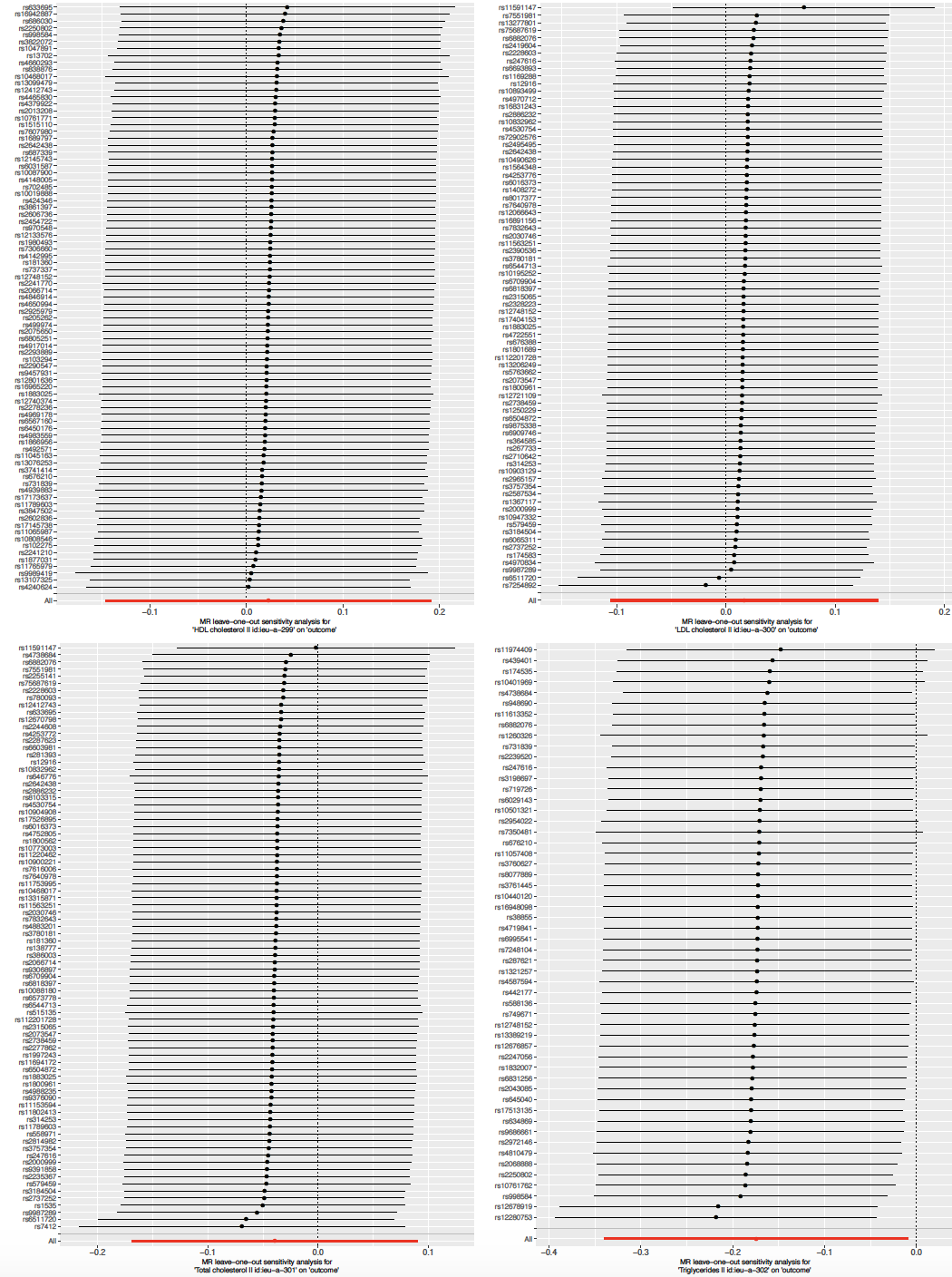

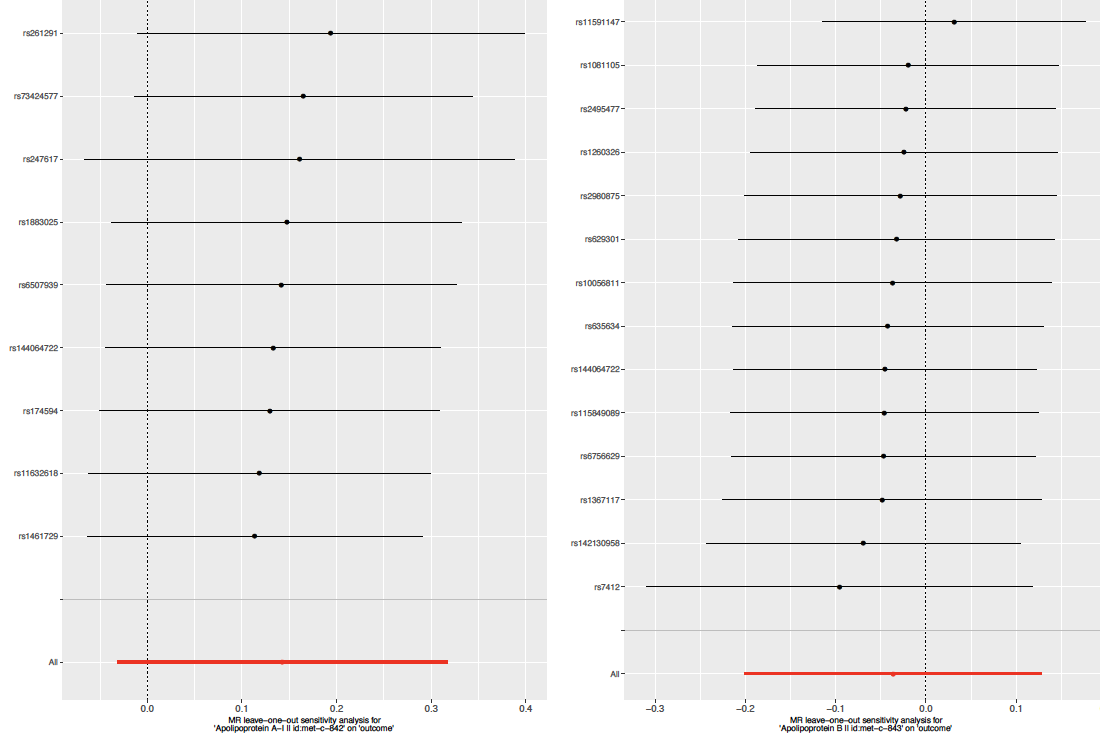

**S7 Fig.** Forest plots showing meta-analysed causal effects of cholesterol-lowering *HMGCR*, *NPC1L1*, *CETP*, *LDLR* and *PCSK9* single nucleotide polymorphisms on combined head and neck cancer in UK Biobank and GAME-ON.

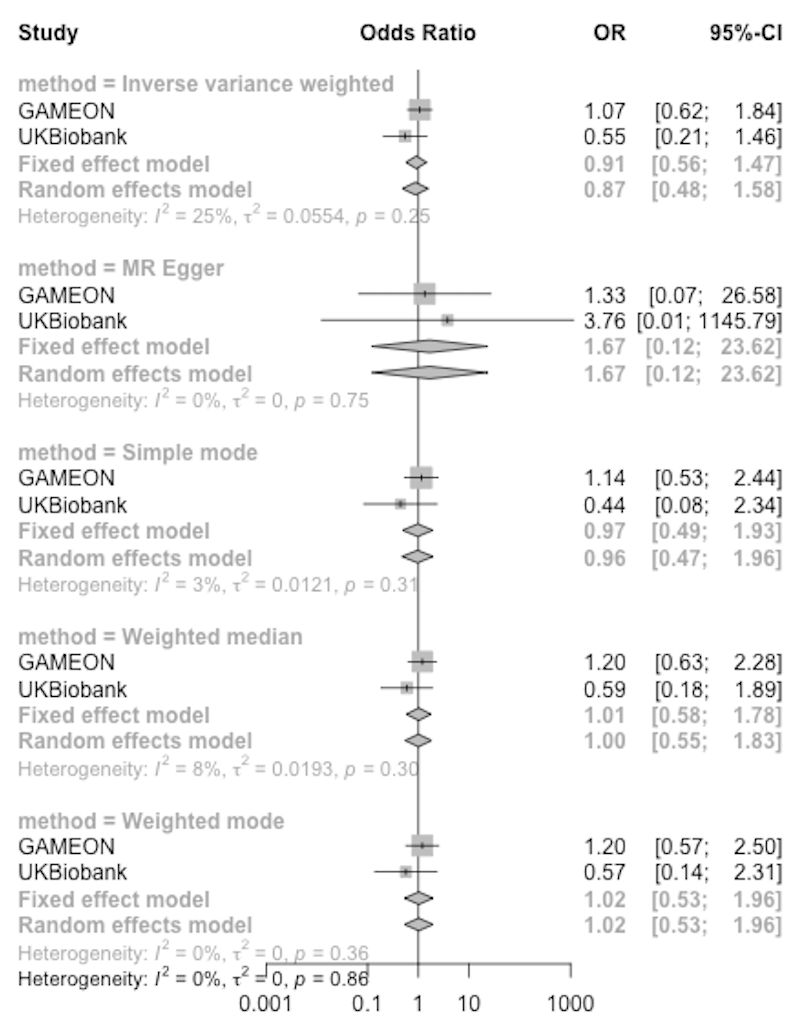
 **HMGCR	NPC1L1**

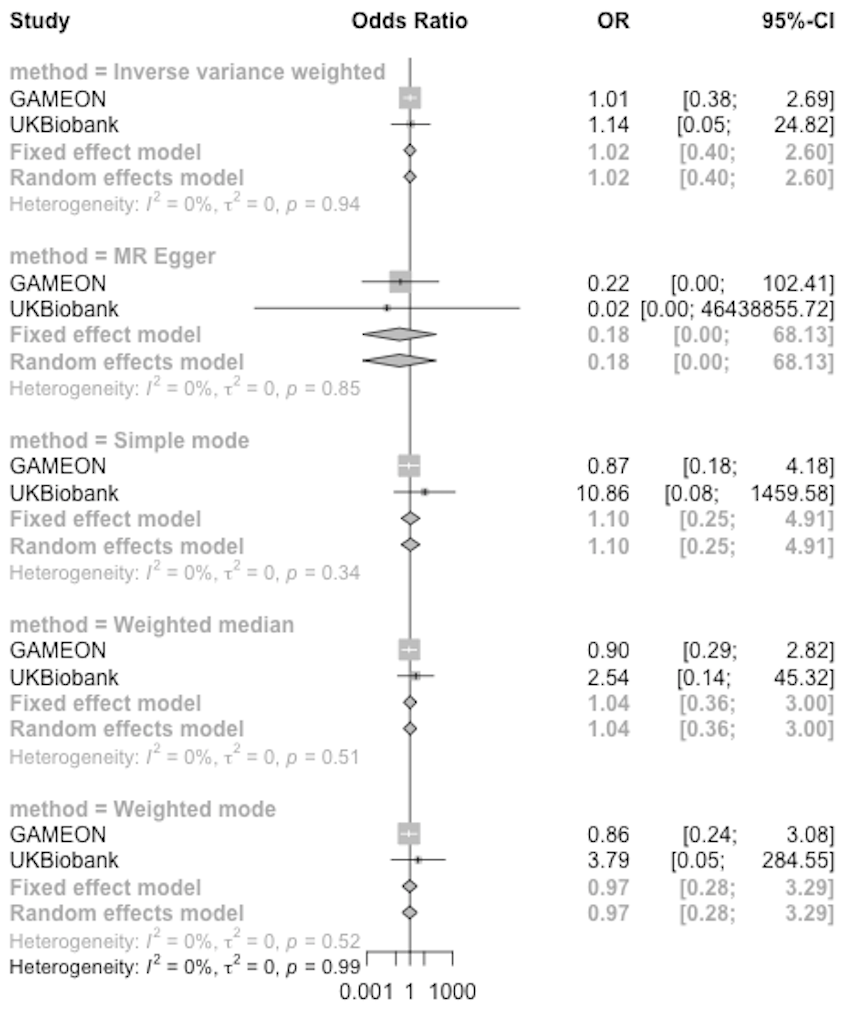

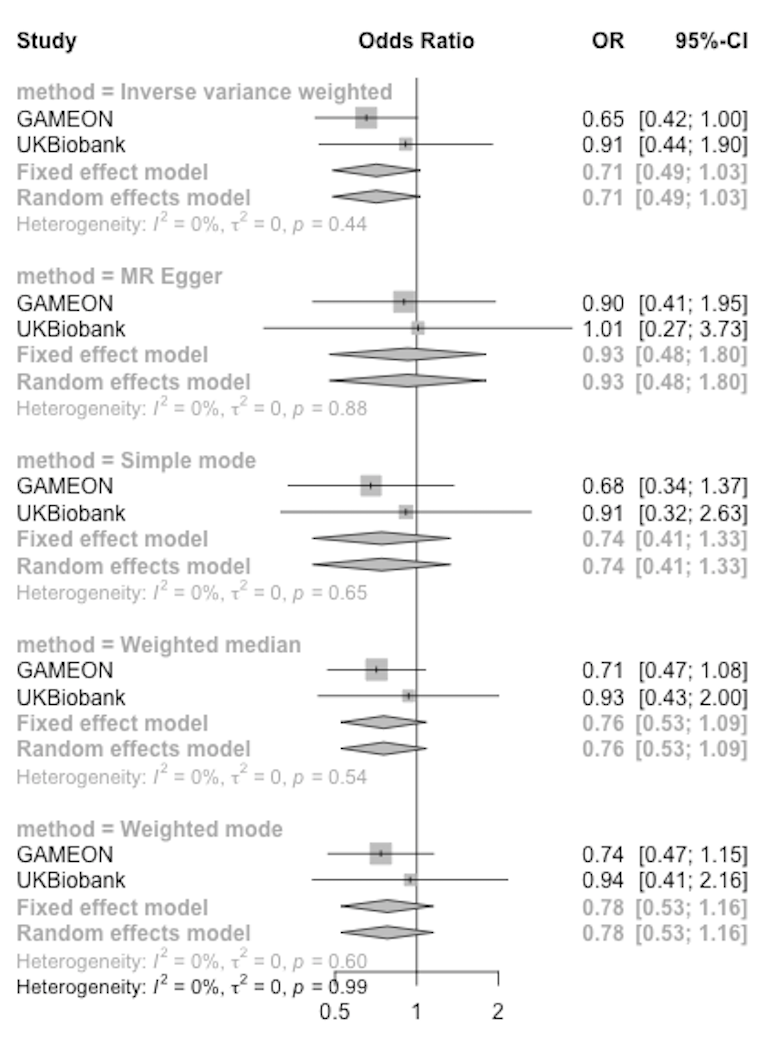

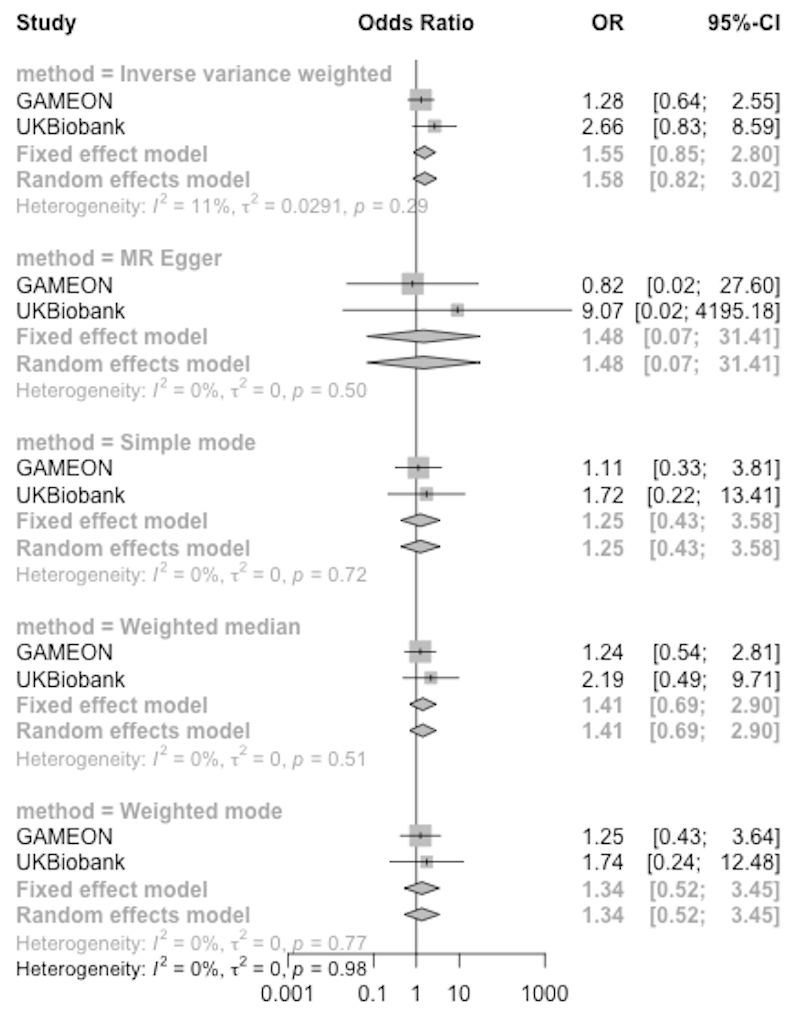
 **CETP	LDLR**

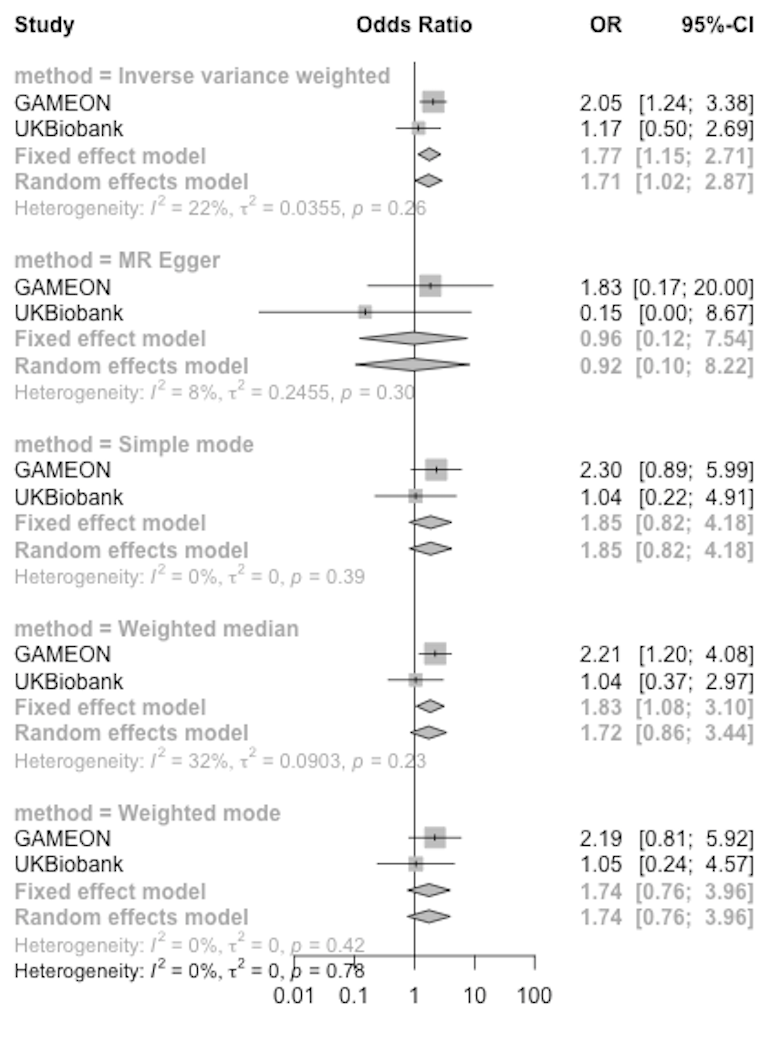
**PCSK9**
