## Supplementary material for "Using genetic variants to evaluate the causal effect of cholesterol lowering on head and neck cancer risk: a Mendelian randomization study": STROBE checklist

**STROBE Statement**

Checklist of items that should be included in reports of observational studies

| **Section/Topic** | Item No | Recommendation | Section/  Page no. (line no.) |
| --- | --- | --- | --- |
| **Title and abstract** | 1 | (*a*) Indicate the study’s design with a commonly used term in the title or the abstract | Title page 1 (1-3) |
|  |  | (*b*) Provide in the abstract an informative and balanced summary of what was done and what was found | Abstract: 5-6 (76-118) |
| Introduction | | | |
| Background/rationale | 2 | Explain the scientific background and rationale for the investigation being reported | Introduction: 8-10 (143-213) |
| Objectives | 3 | State specific objectives, including any prespecified hypotheses | Introduction:  10 (lines 210-213) |
| Methods | | | |
| Study design | 4 | Present key elements of study design early in the paper | Introduction:  10 (lines 194-204) |
| Setting | 5 | Describe the setting, locations, and relevant dates, including periods of recruitment, exposure, follow-up, and data collection | Methods: 11-16 (215-357) |
| Participants | 6 | (*a*) *Cohort study*—Give the eligibility criteria, and the sources and methods of selection of participants. Describe methods of follow-up  *Case-control study*—Give the eligibility criteria, and the sources and methods of case ascertainment and control selection. Give the rationale for the choice of cases and controls  *Cross-sectional study*—Give the eligibility criteria, and the sources and methods of selection of participants | Methods: 11-16 (215-357)  Further information in each of the GWAS publications |
|  |  | (*b*) *Cohort study*—For matched studies, give matching criteria and number of exposed and unexposed  *Case-control study*—For matched studies, give matching criteria and the number of controls per case |  |
| Variables | 7 | Clearly define all outcomes, exposures, predictors, potential confounders, and effect modifiers. Give diagnostic criteria, if applicable | Methods: 11-16 (215-357) |
| Data sources/measurement | 8* | For each variable of interest, give sources of data and details of methods of assessment (measurement). Describe comparability of assessment methods if there is more than one group | Methods: 11-16 (215-357) |
| Bias | 9 | Describe any efforts to address potential sources of bias | Methods: 14-16 (297-324) |
| Study size | 10 | Explain how the study size was arrived at | N/A |
| Quantitative variables | 11 | Explain how quantitative variables were handled in the analyses. If applicable, describe which groupings were chosen and why | Methods: 11-16 (215-357) |
| Statistical methods | 12 | (*a*) Describe all statistical methods, including those used to control for confounding | Methods: 14-16 (297-324) |
|  |  | (*b*) Describe any methods used to examine subgroups and interactions | Methods: 15 (326-330) |
|  |  | (*c*) Explain how missing data were addressed | N/A |
|  |  | (*d*) *Cohort study*—If applicable, explain how loss to follow-up was addressed  *Case-control study*—If applicable, explain how matching of cases and controls was addressed  *Cross-sectional study*—If applicable, describe analytical methods taking account of sampling strategy | N/A |
|  |  | (*e*) Describe any sensitivity analyses | Results: 24 (442-449) |
